# Effect of Intensive vs Standard Blood Pressure Control According to *APOE* ε4 Genotype: A Secondary Analysis of SPRINT

**DOI:** 10.64898/2026.07.02.26357167

**Authors:** Yizhe Xu, Jeremy J. Pruzin, Tom Greene, Jeff Williamson, David M. Reboussin, Nicholas M. Pajewski, Michael Klein, Wai-YingWendy Yau, Eric M. Reiman, Mark A. Supiano, Nicholas Ashton, Jasmeer Chhatwal, Adam P. Bress

**Author notes:** Corresponding Author: Yizhe Xu, PhD, University of Utah Internal Medicine, 295 Chipeta Way, Salt Lake City, Utah 84112-5820.

## Abstract

**Importance:** Apolipoprotein E (*APOE*) ε4 is the strongest genetic risk factor for sporadic dementia, yet whether the benefits of intensive systolic blood pressure (SBP) control differ by *APOE* ε4 carrier status remains unknown. This is among the first randomized evaluations of intensive SBP control on all-cause dementia by *APOE* ε4 status in US adults without diabetes.

**Objective:** To compare effects of intensive vs standard SBP control on incident all-cause probable dementia between *APOE* ε4 carriers and non-carriers.

**Design, Setting, and Participants:** Secondary analysis of the Systolic Blood Pressure Intervention Trial (SPRINT), a multicenter randomized trial of adults ≥50 years with hypertension and increased cardiovascular risk, but without diabetes, prior stroke, or dementia. Primary cognitive follow-up ended July 2018. Participants were further followed with telephone-based outcome assessment from 2019 through 2023.

**Interventions:** Intensive SBP control (goal <120 mm Hg) vs standard SBP control (goal <140 mm Hg).

**Main Outcomes and Measures:** The primary outcome was all-cause probable dementia. Secondary outcomes were mild cognitive impairment (MCI); MCI or dementia; MCI, dementia, or death; and all-cause mortality. *APOE* ε4 status was the primary effect modifier.

**Results:** Of 9,361 randomized participants, 8,390 (89.6%) had *APOE* genotyping (29% ε4 carriers); 7,733 (82.6%) had both genotype and outcome data (mean age, 68 years; 36% female; 28% Non-Hispanic Black). Over a median follow-up of 5.1 years, dementia rates (intensive vs standard) were 9.7 vs 13.2 per 1,000 person-years among ε4 carriers (hazard ratio [HR], 0.73; 95% CI, 0.51-1.04) and 6.1 vs 6.7 among non-carriers (HR, 0.91; 95% CI, 0.67-1.23; P-interaction = .35). Four-year risk differences were −1.7% (95% CI, −3.4% to 0%; number needed to treat, 59) among carriers and 0.2% (95% CI, −0.6% to 1%) among non-carriers (P-interaction = .045). Patterns were similar for the composite of MCI or dementia; effects on MCI were similar between *APOE* ε4 subgroups.

**Conclusions and Relevance:** Among US adults at high cardiovascular risk, intensive SBP control yielded larger absolute risk reduction in dementia among *APOE* ε4 carriers than non-carriers; relative effects were similar. These findings may inform *APOE* ε4-stratified blood pressure management for dementia prevention.

**KEY POINTS:** *Question:* What is the effect of intensive vs standard systolic blood pressure (SBP) control on incident dementia by *APOE* ε4 status?

*Findings:* In SPRINT, intensive vs standard SBP control yielded a significantly larger 4-year absolute risk reduction in dementia among *APOE* ε4 carriers (−1.7%) than non-carriers (0.2%; P-interaction = .045); the relative-scale interactions were directionally consistent but not significant.

*Meaning:* Among US adults at high cardiovascular risk, intensive SBP control yielded larger absolute benefit among *APOE* ε4 carriers than non-carriers; relative benefits were similar. *APOE* ε4 status may inform risk-stratified blood pressure management for dementia prevention.

## INTRODUCTION

Hypertension is among the most prevalent modifiable risk factors for cognitive decline and dementia.^1–5^ Hypertension is associated with accelerated brain aging, greater white matter disease burden, and increased risk of Alzheimer disease (AD) and related dementias.^2–4^ The Systolic Blood Pressure Intervention Trial (SPRINT) demonstrated that intensive systolic blood pressure (SBP) control (<120 mm Hg) compared with standard control (<140 mm Hg) significantly reduced cardiovascular events and all-cause mortality among adults aged 50 years and older with hypertension, a finding that has now been reproduced across at least five additional randomized trials.^6^ SPRINT also showed that intensive SBP control reduced the risk of mild cognitive impairment (MCI) and the combined composite outcome of MCI or dementia, though the reduction in dementia alone did not reach statistical significance.^7–10^

The Apolipoprotein E (*APOE*) ε4 allele is the strongest and most common genetic risk factor for sporadic AD, with approximately 25% of the population carrying at least one copy.^11–13^ Prior meta-analyses and large-scale genetic association studies have reported a 3-4 fold increased risk of AD among *APOE* ε4 heterozygotes and up to 15-fold among homozygotes.^11, 13^ Beyond its well-established role in amyloid-driven neurodegeneration,^14, 15^ ε4 exerts prominent cerebrovascular effects, including accelerated blood-brain barrier breakdown and increased vulnerability to ischemic white matter injury.^16–18^ Observational data indicate that combined hypertension and ε4 carriage synergistically accelerates cognitive decline,^19–23^ suggesting that the cognitive benefits of intensive blood pressure lowering may differ by *APOE* ε4 status.

Despite the established roles of both hypertension and *APOE* ε4 in dementia pathogenesis, no prior randomized trial has examined whether benefits of intensive SBP control on dementia risk differ by ε4 carrier status. Using data from SPRINT, we compared the effect of intensive vs standard SBP control on incident dementia, MCI, and their composite between ε4 carriers and non-carriers, assessing heterogeneity on both relative and absolute scales. We further leveraged extended follow-up from SPRINT to examine the durability of these findings and assessed risk profiles by ε4 and ε2 carrier status and allele count.

## METHODS

### Study Approval, Trial Design and Study Participants

This study was approved by the Institutional Review Boards of the University of Utah. The design and primary results of SPRINT have been described in detail previously.^24, 25^ Briefly, SPRINT was a multicenter, open-label, randomized clinical trial conducted at 102 sites in the United States and Puerto Rico. Between November 2010 and March 2013, 9,361 adults aged 50 years or older with SBP of 130 mm Hg or higher and increased cardiovascular risk, but without diabetes mellitus, prior stroke, or dementia, were randomized 1:1 to intensive (SBP <120 mm Hg) or standard (SBP <140 mm Hg) blood pressure control. The trial was approved by institutional review boards at each site, and all participants provided written informed consent (ClinicalTrials.gov: NCT01206062).

### Outcomes

Detailed methods for cognitive assessment and outcome adjudication have been reported previously,^8, 9^ and are summarized in the Supplemental Methods. The primary outcome was incident dementia. Secondary outcomes included MCI; the composite of MCI or dementia; the composite of MCI, dementia, or death; and all-cause mortality. MCI required two consecutive adjudicated classifications of MCI (eFigure 2).

### Study Follow-up

The blood pressure intervention was stopped early on August 20, 2015 (median intervention period, 3.26 years) after an interim analysis showed that the primary composite cardiovascular endpoint and all-cause mortality favored intensive treatment. ^24^ Cognitive status was assessed during the trial and across two subsequent extensions of follow-up. In-person cognitive evaluations were planned at baseline, 2 years, 4 years, and a closeout visit conducted through July 2018, though early termination of the intervention introduced some variability in the timing of the later assessments (median follow-up, 5.1 years).^9, 24^ A second, telephone-based extension ascertained cognitive status from December 2019 through December 2023, extending the median follow-up to 6.9 years and accruing an additional 216 adjudicated dementia cases (eFigure 1).^8^ We used the cognitive follow-up through July 2018 as our primary analysis period and the extended follow-up through December 2023 as a secondary analysis period.

### *APOE* **ε**4 Genotyping and Effect Modifier Definition

*APOE* genotyping was performed as a novel addition to SPRINT, leveraging stored DNA samples collected and maintained at the University of Utah. Genotyping was conducted at the Utah Genomics Core using TaqMan allelic discrimination assays for rs429358 and rs7412.^26^ Diplotype-to-ε4 allele coding is provided in the Supplemental Methods. In our primary analyses, participants were classified as *APOE* ε4 carriers (ε4 count ≥1) or ε4 non-carriers (ε4 count = 0), which served as the primary effect modifier for all treatment comparisons. In secondary analyses, we further stratified participants by ε4 allele count, as ε4 non-carriers (0), ε4 heterozygotes (1), and ε4 homozygotes (2). ε2 carrier status, ε2 allele count, and a mutually exclusive ε2/ε3/ε4 classification (ε2/ε2 or ε2/ε3 as ε2 carriers; ε3/ε3 as ε3; any ε4 allele as ε4 carriers) were defined analogously.

### Baseline Measures

We preselected 27 established risk factors for cognitive decline and assessed baseline balance between treatment groups within each *APOE* ε4 subgroup. Covariates spanned age, sex, race/ethnicity, education, insurance, systolic and diastolic blood pressure, estimated glomerular filtration rate, fasting glucose, total cholesterol, high-density lipoprotein cholesterol, body mass index, smoking status, statin use, aspirin use, family history of heart disease, Framingham score, and antihypertensive medication use by class (angiotensin-converting enzyme inhibitors or angiotensin receptor blockers, beta-blockers, calcium channel blockers, thiazide diuretics, loop diuretics, alpha-blockers, and other antihypertensive agents), and baseline cognitive performance measures including the Montreal Cognitive Assessment (MoCA), Digit Symbol Coding Test, and Logical Memory Delayed Recall.

### Statistical Analysis

Using the primary trial follow-up period, we fit separate Cox proportional hazards models for *APOE* ε4 carriers and non-carriers to estimate cause-specific hazard ratios (HRs) with 95% confidence intervals (CIs), with participants who died prior to developing dementia censored at the time of death. To complement HRs, we estimated 4-year risk differences (RDs) and risk ratios (RRs) using the Aalen-Johansen estimator^27^ within a competing risks framework, where we estimated the cumulative incidence function (CIF) of dementia and dementia-free death as competing events (Supplemental Methods). We computed the 95% CIs of RDs and RRs using the delta method applied to the pointwise variance of treatment-specific CIFs estimated on the Wald scale.^28, 29^ We also computed the number needed to treat (NNT) as the inverse of RD. Heterogeneity of treatment effects by *APOE* ε4 carrier status was assessed using Wald tests on the HR, RD, and RR scales. The proportional hazards assumption was evaluated using Schoenfeld residual tests.

We conducted four secondary analyses. First, all primary analyses were repeated for secondary outcomes. Second, treatment effects were compared across ε2, ε3, and ε4 carriers. Third, to evaluate the independent association between *APOE* ε4 genotype and risk of MCI and dementia in the full cohort, we fit Cox models under two genetic frameworks: a dominant model (carrier vs non-carrier) and an allelic count model (0, 1, or 2 ε4 alleles). The allelic count was modeled as a continuous variable, assuming a constant hazard ratio per additional ε4 allele. To assess the extent to which demographic factors attenuated the genotype-outcome association, we also fit a model adjusted for age, sex, and race/ethnicity. Fourth, we repeated the association analysis above with *APOE* ε*2* genotype as the exposure. In sensitivity analyses, all primary and secondary analyses were repeated using the extended follow-up from SPRINT to assess the durability of findings. All analyses were performed using R version 4.4.0 (R Foundation for Statistical Computing).

## RESULTS

### Study Participants

Of the 9,361 randomized SPRINT participants, 7,733 (83%) had non-missing *APOE* ε4 genotype and clinical outcome data and were included in the present study (Figure 1). The mean age was 68 years (SD, 9.3), 36% were female, 28% were Non-Hispanic Black, and 11% were Hispanic. Baseline characteristics were similar to those of the full trial cohort (eTable 2). At 12 months, the mean between-group difference in achieved SBP was −15 mm Hg among ε4 carriers and non-carriers (eFigure 3), indicating that the blood pressure separation achieved by randomization was comparable across genotype subgroups.

**Figure 1.**
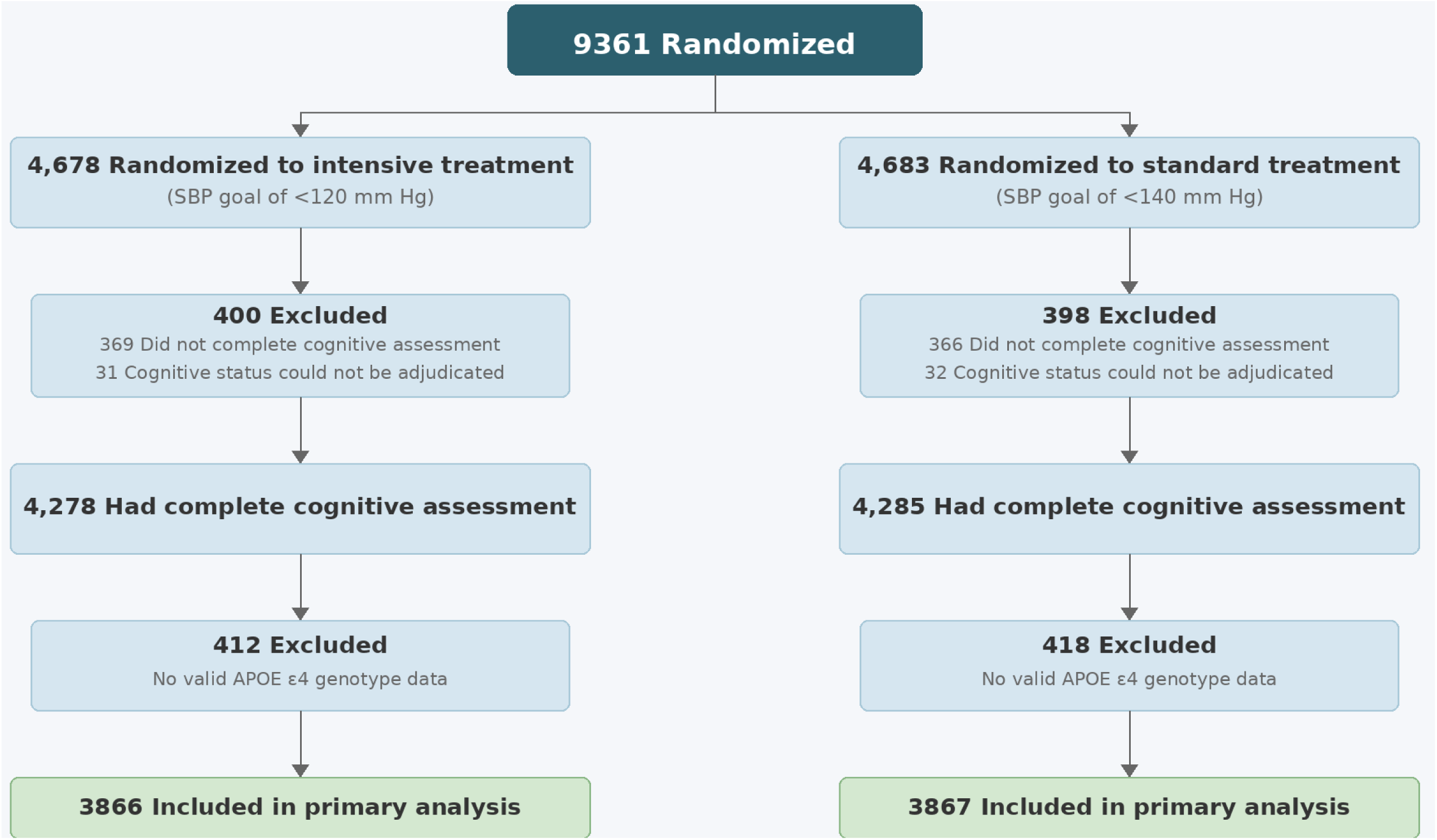
Randomization and Follow-Up for MCI and Dementia Outcomes in the Systolic Blood Pressure Intervention Trial. Participants with missing APOE genotype or clinical outcomes were excluded.

### *APOE* **ε**4 Genotyping and Carrier Status

Among the 9,361 SPRINT participants, 8,451 stored DNA samples from the University of Utah were genotyped, yielding 8,390 *APOE* ε4 determinations (61 [<1%] undetermined). Of these, 5,957 (71%) were non-carriers, 2,219 (26%) were heterozygous, and 214 (2.5%) were homozygous. The distribution was similar among the 7,733 participants included in the present analysis: 5,492 (71%) non-carriers, 2,049 (26%) heterozygous, and 192 (2.5%) homozygous (eTable 2). Non-Hispanic Black participants had a higher prevalence of ε4 heterozygosity (35%) and homozygosity (4.6%) compared with the overall cohort. *APOE* ε4 genotype distributions were consistent with Hardy-Weinberg equilibrium in the overall sample and within each race/ethnicity stratum (all P ≥ .15) (eTable 1).

### Primary Outcome

Baseline characteristics were similar between intensive and standard treatment groups within both ε4 carriers and non-carriers (Table 1). During the primary trial follow-up, dementia was adjudicated in 82 participants (6.1 per 1,000 person-years) in the intensive treatment group vs 88 (6.7 per 1,000 person-years) in the standard treatment group among ε4 non-carriers (HR, 0.91; 95% CI, 0.67-1.23), and in 51 participants (9.7 per 1,000 person-years) vs 72 (13.2 per 1,000 person-years) among ε4 carriers (HR, 0.73; 95% CI, 0.51-1.04) (Figure 2 and Figure 3). Test of treatment-by-ε4 interaction on HRs did not reach statistical significance (P-interaction = .35) and was materially unchanged after adjustment for age, baseline MoCA, or both (eTable 17).

**Table 1.** Baseline Characteristics by Randomized Treatment Group and *APOE* ε*4* Carrier Status.

| Characteristic | Non-carriers |  |  | Carriers |  |  |
| --- | --- | --- | --- | --- | --- | --- |
|  | Standard<br>N = 2,711<sup>1</sup> | Intensive<br>N = 2,781<sup>1</sup> | SMD<sup>2</sup> | Standard<br>N = 1,156<sup>1</sup> | Intensive<br>N = 1,085<sup>1</sup> | SMD<sup>2</sup> |
| **Age** | 68.5 (9.4) | 68.5 (9.3) | 0.01 | 66.6 (9.2) | 66.8 (9.0) | -0.02 |
| **Female** | 959 (35.4%) | 988 (35.5%) | -0.15% | 429 (37.1%) | 403 (37.1%) | -0.03% |
| **Race** |  |  | 0.03 |  |  | 0.12 |
| NH White | 1,723 (63.6%) | 1,742 (62.6%) |  | 584 (50.5%) | 568 (52.4%) |  |
| NH Black | 641 (23.6%) | 663 (23.8%) |  | 452 (39.1%) | 399 (36.8%) |  |
| Hispanic | 300 (11.1%) | 318 (11.4%) |  | 109 (9.4%) | 93 (8.6%) |  |
| Other | 47 (1.7%) | 58 (2.1%) |  | 11 (1.0%) | 25 (2.3%) |  |
| **Education Level** |  |  | 0.04 |  |  | 0.05 |
| College graduate | 1,137 (41.9%) | 1,168 (42.0%) |  | 434 (37.5%) | 397 (36.6%) |  |
| High school diploma or GED | 409 (15.1%) | 439 (15.8%) |  | 181 (15.7%) | 189 (17.4%) |  |
| Less than high school diploma | 252 (9.3%) | 230 (8.3%) |  | 110 (9.5%) | 96 (8.8%) |  |
| Some college (No degree) | 913 (33.7%) | 944 (33.9%) |  | 431 (37.3%) | 403 (37.1%) |  |
| **Private Insurance** | 1,203 (44.4%) | 1,246 (44.8%) | -0.43% | 484 (41.9%) | 483 (44.5%) | -2.6% |
| **Systolic Blood Pressure** | 140.2 (15.3) | 139.7 (15.8) | 0.03 | 139.2 (15.3) | 139.7 (16.2) | -0.03 |
| **Diastolic Blood Pressure** | 77.8 (12.0) | 77.8 (11.6) | -0.01 | 78.8 (11.8) | 78.9 (12.2) | -0.01 |
| **eGFR** | 71.1 (20.1) | 71.4 (20.2) | -0.02 | 73.1 (21.1) | 72.6 (21.0) | 0.03 |
| **Glucose** | 99.2 (13.3) | 99.0 (13.7) | 0.02 | 97.5 (13.6) | 98.8 (13.6) | -0.09 |
| **Total Cholesterol** | 188.0 (40.2) | 187.9 (41.2) | 0.00 | 195.4 (41.3) | 194.8 (41.7) | 0.01 |
|  | Standard<br>N = 2,711<sup>1</sup> | Intensive<br>N = 2,781<sup>1</sup> | SMD<sup>2</sup> | Standard<br>N = 1,156<sup>1</sup> | Intensive<br>N = 1,085<sup>1</sup> | SMD<sup>2</sup> |
| **HDL Cholesterol** | 53.0 (14.5) | 53.1 (14.3) | -0.01 | 52.4 (14.7) | 51.8 (13.9) | 0.04 |
| **Statin Use** | 1,168 (43.1%) | 1,174 (42.2%) | 0.87% | 561 (48.5%) | 492 (45.3%) | 3.2% |
| **Aspirin Use** | 1,410 (52.0%) | 1,513 (54.4%) | -2.4% | 604 (52.2%) | 576 (53.1%) | -0.84% |
| **Smoking Status** |  |  | 0.02 |  |  | 0.01 |
| Current | 303 (11.2%) | 327 (11.8%) |  | 167 (14.5%) | 157 (14.5%) |  |
| Former | 1,182 (43.7%) | 1,200 (43.2%) |  | 480 (41.6%) | 446 (41.1%) |  |
| Never | 1,222 (45.1%) | 1,253 (45.1%) |  | 508 (44.0%) | 482 (44.4%) |  |
| **Body Mass Index** | 29.7 (5.6) | 29.9 (5.8) | -0.03 | 29.9 (6.0) | 29.9 (5.7) | 0.00 |
| **ACEI or ARB** | 1,607 (59.3%) | 1,662 (59.8%) | -0.49% | 626 (54.2%) | 633 (58.3%) | -4.2% |
| **Beta Blocker** | 940 (34.7%) | 1,045 (37.6%) | -2.9% | 399 (34.5%) | 421 (38.8%) | -4.3% |
| **Loop Diuretic** | 143 (5.3%) | 163 (5.9%) | -0.59% | 70 (6.1%) | 72 (6.6%) | -0.58% |
| **Calcium Channel Blocker** | 948 (35.0%) | 958 (34.4%) | 0.52% | 402 (34.8%) | 372 (34.3%) | 0.49% |
| **Thiazide Diuretic** | 1,122 (41.4%) | 1,088 (39.1%) | 2.3% | 465 (40.2%) | 440 (40.6%) | -0.33% |
| **Alpha Blocker** | 272 (10.0%) | 303 (10.9%) | -0.86% | 110 (9.5%) | 90 (8.3%) | 1.2% |
| **Other Antihypertensive Medication** | 92 (3.4%) | 109 (3.9%) | -0.53% | 51 (4.4%) | 47 (4.3%) | 0.08% |
| **Family History of Heart Disease** |  |  | 0.04 |  |  | 0.07 |
| No | 883 (32.6%) | 859 (30.9%) |  | 412 (35.6%) | 356 (32.8%) |  |
| Unknown | 156 (5.8%) | 153 (5.5%) |  | 61 (5.3%) | 53 (4.9%) |  |
| Yes | 1,668 (61.6%) | 1,769 (63.6%) |  | 683 (59.1%) | 676 (62.3%) |  |
|  | Standard<br>N = 2,711<sup>1</sup> | Intensive<br>N = 2,781<sup>1</sup> | SMD<sup>2</sup> | Standard<br>N = 1,156<sup>1</sup> | Intensive<br>N = 1,085<sup>1</sup> | SMD<sup>2</sup> |
| **Framingham Risk Score** | 22.3 (15.4, 32.3) | 22.1 (15.3, 31.8) | 0.02 | 21.6 (14.9, 31.3) | 21.9 (15.3, 31.5) | -0.06 |
| **MoCA Score** | 24.0 (21.0, 26.0) | 24.0 (21.0, 26.0) | 0.01 | 23.0 (20.0, 26.0) | 23.0 (21.0, 26.0) | -0.04 |
| **Digit Symbol Coding Test** | 51.6 (15.2) | 51.6 (15.4) | -0.01 | 51.0 (15.6) | 51.0 (14.6) | 0.00 |
| **Logical Memory Delayed Recall** | 8.4 (3.3) | 8.4 (3.3) | 0.02 | 8.0 (3.5) | 8.3 (3.3) | -0.08 |

<sup>1</sup>Mean (SD); n (%); Median (Q1, Q3)
<sup>2</sup>Standardized Mean Difference; 2-sample test for equality of proportions with continuity correction

**Figure 2.**
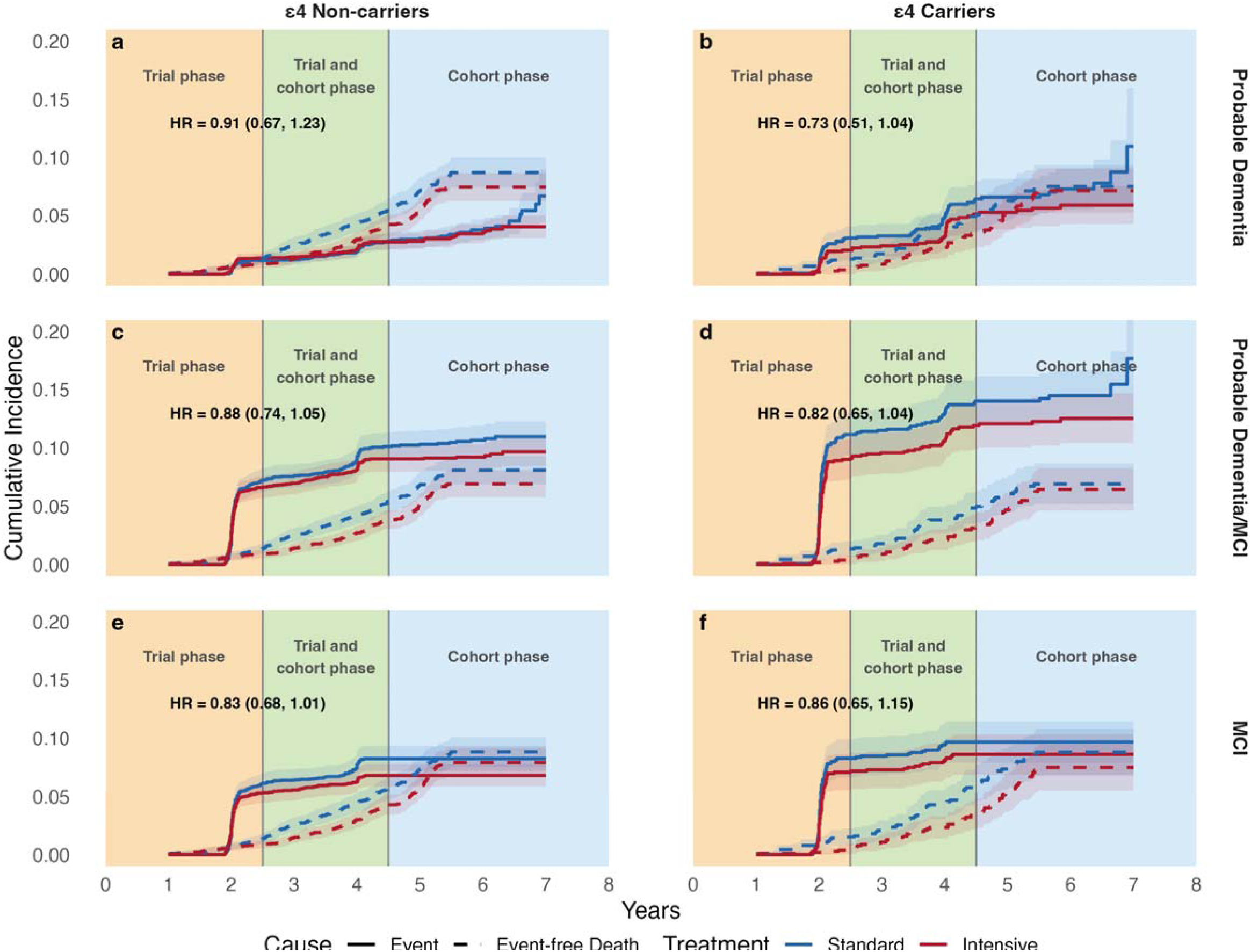
Cumulative incidence curves by *APOE* ε*4* carrier status for MCI and dementia outcomes. Cumulative incidence curves show the risk of event and event-free death under the intensive vs standard SBP control for *APOE* ε*4* carriers and non-carriers, estimated using a competing risk framework. Shaded areas indicate 95% confidence intervals.

During the primary follow-up, 293 participants (3.8%) developed dementia and 444 (5.7%) died without developing dementia, with competing deaths exceeding the primary outcome of interest. In the competing risks analysis, intensive SBP control yielded a greater reduction at 4 years in dementia among *APOE* ε4 carriers (RD, −1.7%; 95% CI, −3.4% to 0.0%; RR, 0.65; 95% CI, 0.43-1.00; NNT = 59) than among non-carriers (RD, 0.2%; 95% CI, −0.6% to 1.0%; RR, 1.10; 95% CI, 0.75-1.60), and significantly reduced the risk of dementia-free death in both subgroups (Table 2; eFigure 5). The treatment-by-ε4 interaction was statistically significant on the RD scale but not on the RR scale (P-interaction = .045 for RD; P-interaction = .07 for RR).

**Table 2.** Assessing treatment effect heterogeneity by. ε**4 carrier status using a competing risk framework.**

| Outcome<sub>3</sub> | ε4 non-carriers |  |  |  | ε4 Carriers |  |  |  | P for interaction on RD<sup>2</sup> | P for interaction on RR |
| --- | --- | --- | --- | --- | --- | --- | --- | --- | --- | --- |
|  | 4-year Risk |  | RD<sup>1</sup><br>(95% CI) | RR<br>(95% CI) | 4-year Risk |  | RD<br>(95% CI) | RR<br>(95% CI) |  |  |
|  | Standard | Intensive |  |  | Standard | Intensive |  |  |  |  |
| Primary Outcome |  |  |  |  |  |  |  |  |  |  |
| Dementia | 0.020 | 0.022 | 0.002<br>(-0.006, 0.010) | 1.10<br>(0.75, 1.60) | 0.050 | 0.032 | -0.017<br>(-0.034, -0.000) | 0.65<br>(0.43, 1.00) | 0.045 | 0.07 |
| Dementia-free death | 0.044 | 0.030 | -0.014<br>(-0.024, -0.004) | 0.68<br>(0.51, 0.91) | 0.039 | 0.023 | -0.016<br>(-0.031, -0.001) | 0.59<br>(0.36, 0.97) | -- | -- |
| Secondary Outcomes |  |  |  |  |  |  |  |  |  |  |
| Dementia or MCI | 0.093 | 0.082 | -0.011<br>(-0.026, 0.004) | 0.88<br>(0.74, 1.05) | 0.131 | 0.107 | -0.024<br>(-0.051, 0.003) | 0.82<br>(0.65, 1.03) | 0.41 | 0.60 |
| Dementia or MCI-free death | 0.042 | 0.027 | -0.015<br>(-0.025, -0.005) | 0.64<br>(0.47, 0.86) | 0.038 | 0.021 | -0.017<br>(-0.032, -0.003) | 0.55<br>(0.33, 0.92) | -- | -- |
| MCI | 0.078 | 0.062 | -0.015<br>(-0.029, -0.001) | 0.80<br>(0.66, 0.98) | 0.096 | 0.082 | -0.014<br>(-0.038, 0.011) | 0.86<br>(0.65, 1.13) | 0.92 | 0.72 |
| MCI-free death | 0.044 | 0.03 | -0.014<br>(-0.025, -0.004) | 0.68<br>(0.51, 0.90) | 0.043 | 0.023 | -0.020<br>(-0.035, -0.004) | 0.55<br>(0.33, 0.90) | -- | -- |

<sup>1</sup>CI = confidence interval; RD = risk difference; RR = risk ratio; MCI = mild cognitive impairment. RRs and RDs compare intensive SBP control (<120 mm Hg) with standard SBP control (<140 mm Hg) as referent.
<sup>2</sup>Interaction p-values are from Wald tests of the treatment × APOE $\epsilon 4$ interaction term.
<sup>3</sup>This analysis uses the primary SPRINT follow-up period without adjusting for baseline covariates.

### Secondary Outcomes

During the primary follow-up, incidence rates per 1,000 person-years (intensive vs standard) were 18.5 vs 21.0 for the composite of MCI or dementia among ε4 non-carriers (242 vs 265 events) and 24.6 vs 30.2 among ε4 carriers (124 vs 157 events); for MCI alone, rates were 13.6 vs 16.6 among non-carriers (178 vs 208 events) and 17.4 vs 20.4 among carriers (87 vs 105 events). There was no interaction by ε4 carrier status (P-interaction = .60) (Figure 3). However, we observed statistically larger protective effects on the composite of MCI or dementia among ε2 carriers (HR, 0.55; 95% CI, 0.36-0.82) than among ε4 carriers (HR, 0.82; 95% CI, 0.65-1.04) (P-interaction = .03) (eTable 3). For the composite of MCI, dementia, or death, incidence was lower with intensive than standard SBP control in both ε4 carriers (HR, 0.77; 95% CI, 0.63-0.94) and non-carriers (HR, 0.86; 95% CI, 0.75-0.99), with no statistically significant interaction by carrier status (eTable 3; Figure 3).

**Figure 3.**
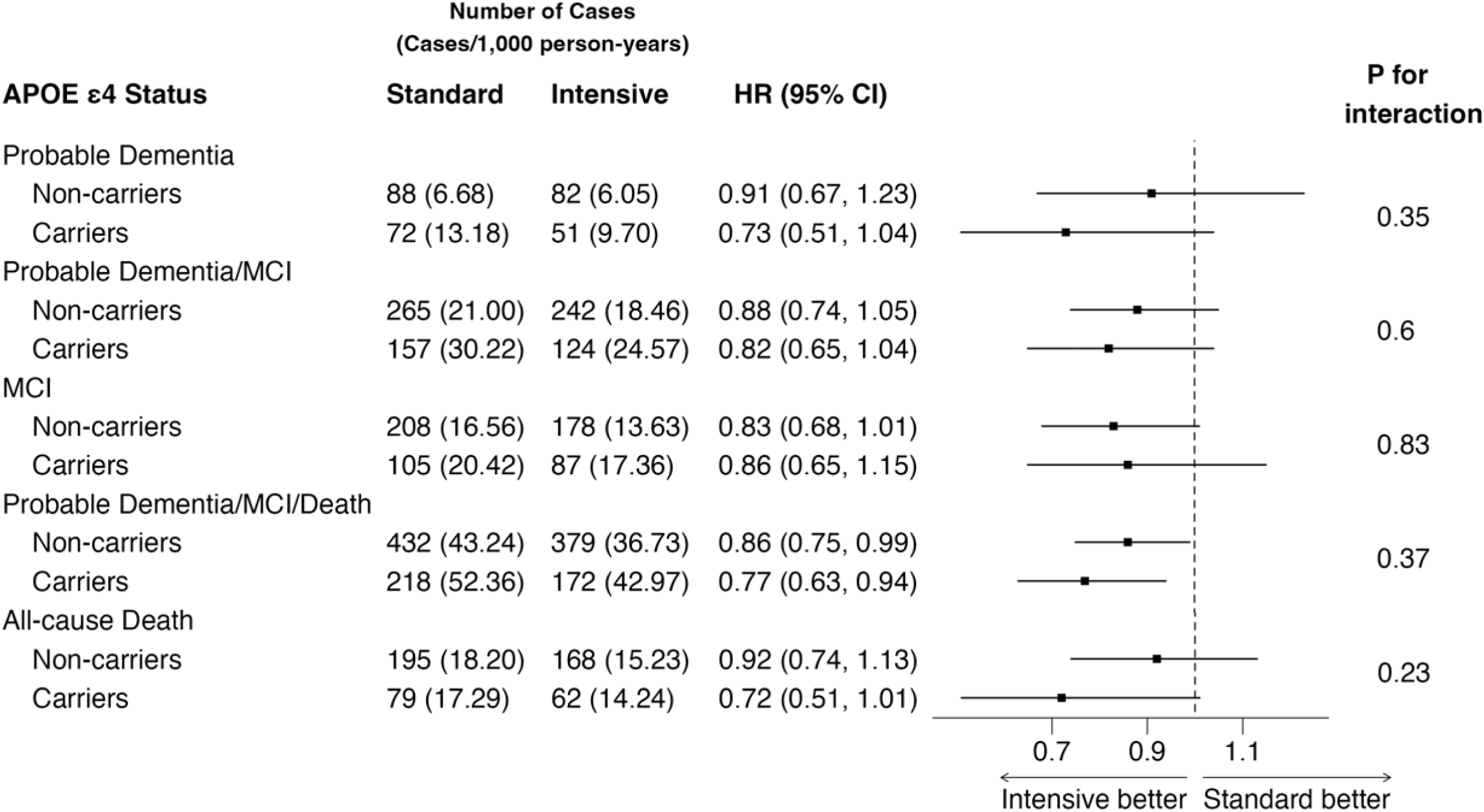
Effects of Intensive vs Standard SBP Control on MCI and Dementia Outcomes by *APOE* ε*4* Carrier Status. CI = confidence interval; HR = hazard ratio; MCI = mild cognitive impairment. Hazard ratios compare intensive SBP control (<120 mm Hg) with standard SBP control (<140 mm Hg) as referent. Interaction p-values are from Wald tests of the treatment × *APOE* ε*4* interaction term. Event rates are per 1000 person-years; mortality rates are reported alongside event rates to assess the impact of the competing event of death. Primary analyses use the primary SPRINT follow-up period without adjusting for baseline covariates. Death events prior to dementia or MCI are censored at the time of death.

### Association of *APOE* **ε**4 Genotype with Clinical Outcomes

In the unadjusted dominant model, *APOE* ε4 carrier status was associated with a higher risk of dementia compared with non-carriers (HR, 1.82; 95% CI, 1.44-2.29) (eTable 4). Significant but attenuated associations were observed between ε4 carrier status and the composite of MCI or dementia, MCI (eTable 4), as well as the composite of MCI, dementia, or death (eTable 5). No association was observed between ε4 genotype and all-cause mortality (eTable 5). Results were similar under the allelic count model across all outcomes (eTables 6-7).

### Association of *APOE* **ε***2* Genotype with Clinical Outcomes

In the unadjusted model, *APOE* ε2 carrier status was associated with a lower risk of dementia compared with non-carriers (HR, 0.66; 95% CI, 0.46-0.95) (eTable 8). No associations were observed between ε2 carrier status and all the secondary outcomes (eTables 8-9). Results were similar under the allelic count model across all outcomes (eTables 10-11).

### Sensitivity Analyses

Under extended follow-up and a competing risk framework (eFigure 4), the 6-year risk of dementia was lower under intensive treatment among ε4 carriers (0.054 vs 0.064; RD, −1.0%; 95% CI, −3.0% to 1.1%; RR, 0.85; 95% CI, 0.60-1.20) but the same among ε4 non-carriers (0.032 vs 0.032; RD, 0%; 95% CI, −1.0% to 0.9%; RR, 0.99; 95% CI, 0.73-1.34) (eTable 12). Treatment effects on dementia-free death at 6 years were not significant for both subgroups (RR: 0.87; 95% CI, 0.62-1.20 for ε4 carriers; RR: 0.84; 95% CI, 0.70-1.02 for ε4 non-carriers). Findings were similar for the composite of MCI or dementia (eTable 12). HR estimates are shown in eFigure 6.

## DISCUSSION

In this secondary analysis of SPRINT, intensive compared with standard SBP control was associated with a larger absolute reduction in adjudicated all-cause dementia among *APOE* ε4 carriers than non-carriers. Effects on the relative risk scale were also directionally consistent with greater relative benefit in carriers but did not differ significantly between groups. Independently of treatment assignment, ε4 carriage was associated with higher dementia risk and ε2 carriage with lower risk. To our knowledge, this is among the first randomized evidence to demonstrate that an intensive BP intervention reduces risk of dementia in cognitively unimpaired *APOE* ε4 carriers, the population at greatest genetic risk for AD.^11–13^

The clinical and public health implications are amplified by the nature of the intervention. Intensive SBP control is inexpensive, broadly available, and immediately actionable with generic antihypertensives, and the 2025 AHA/ACC guideline endorses a goal of <130 mm Hg, with encouragement toward <120 mm Hg, for adults at elevated risk.^30^ The scale of the opportunity is substantial: nearly half of US adults, approximately 120 million, have hypertension, only about 1 in 4 are controlled to the guideline goal, and an estimated 25% to 29% carry at least one ε4 allele.^31, 32^ Among these roughly 75 million adults over age 50, ε4 carriers number^33^ approximately 19 to 22 million; applying the observed 4-year number needed to treat of 59, intensive control could prevent or delay on the order of 320,000 to 370,000 dementia cases over 4 years. Because absolute cognitive benefit scales with baseline risk,^33^ this is an illustrative upper bound that assumes the SPRINT-observed treatment effect across the hypertensive population; realized absolute benefit would be smaller in lower-risk adults, yet even a fraction of this total would represent a substantial public health benefit in a population with few proven prevention options.

Next, these findings reinforce the growing recognition that AD pathology and cerebrovascular disease are closely intertwined in the genesis of cognitive decline. The ε4 isoform confers heightened cerebrovascular vulnerability, accelerating blood-brain barrier breakdown, reducing pericyte coverage, and increasing susceptibility to ischemic white matter injury,^34, 35^ likely rendering the brains of carriers disproportionately sensitive to blood pressure-mediated injury. Observational data further show that hypertension and ε4 carriage synergistically accelerate amyloid deposition and cognitive decline,^36, 37^ and emerging evidence indicates that elevated SBP worsens tau accumulation in the presence of amyloid.^38–40^ The largest absolute benefit emerged in the subgroup with both the greatest AD-related and vascular vulnerability, consistent with intensive SBP control preferentially removing a modifiable vascular driver of dementia in those at highest baseline risk.

This AD-vascular pathology interplay has direct therapeutic implications in the era of amyloid-targeted therapy. These agents are now being evaluated as preventative therapies,^41, 42^ yet they address only one of the two pathologies that most often co-occur in dementia. A combined treatment paradigm, pairing amyloid-targeted therapy with intensive SBP control, merits prospective evaluation both in symptomatic AD, where combined treatment may improve therapeutic safety^43^ and augment disease modification, ^38–40^ and as a primary prevention strategy in cognitively unimpaired but high-risk ε4 carriers.

A key unresolved question is whether this differential benefit reflects an effect on Alzheimer disease pathology, on cerebral amyloid angiopathy (more common in ε4 carriers), on other cerebrovascular changes, or on their interaction. The present analysis cannot differentiate among these, as molecular and imaging markers of these pathways were unavailable. Planned work within the NIH-funded SPRINT VITAL project will address this on two fronts: whether the treatment effect differs by baseline AD pathology, determined by plasma pTau217 at baseline, and whether intensive control alters downstream markers across these pathways, including pTau217 and GFAP (Alzheimer and astroglial), placental growth factor and small-vessel disease on neuroimaging (vascular), and neurofilament light chain (neurodegeneration).

The distinction between relative and absolute treatment effects is central to interpreting these findings. Because ε4 carriers have substantially higher baseline dementia risk than non-carriers, even comparable relative reductions translate into meaningfully larger absolute benefits, reflected in the significantly larger 4-year risk difference among carriers than non-carriers. This is a well-recognized phenomenon in cardiovascular prevention with direct implications for clinical decision-making.^44^ Approximately 25% of the general population carries at least one ε4 allele, with substantially higher prevalence among Non-Hispanic Black individuals, a group already disproportionately burdened by both hypertension and dementia.

This analysis has several strengths: newly available *APOE* genotyping; adjudicated MCI and dementia; assessment across both the primary and extended follow-up periods; and a competing-risks framework jointly modeling dementia and dementia-free death, essential because dementia-free deaths (5.7%) exceeded primary dementia events (3.8%). Several limitations warrant consideration. The study was underpowered to detect treatment-effect heterogeneity, and early termination of the intervention compounded this by narrowing the SBP separation and limiting dementia events. We did not adjust for multiple comparisons. Baseline MCI was not adjudicated, so prevalent MCI at randomization, possibly more common among ε4 carriers, cannot be excluded; screening thresholds may also have underestimated MCI, though likely similarly across groups given blinded adjudication. Limitations of telephone-based outcome assessment have been discussed previously.^8, 9^

## CONCLUSION

In this secondary analysis of SPRINT, the effect of intensive versus standard SBP control on all-cause dementia differed by *APOE* ε4 status on the absolute scale, with a larger absolute risk reduction among carriers, although the relative-scale interaction was not significant. *APOE* ε4 carriage was independently associated with higher dementia risk and *APOE* ε2 carriage with lower risk, both robust to covariate adjustment. Together, these findings may inform *APOE* ε4-stratified blood pressure management for dementia prevention.

## Supporting information

Supplementary Online Content

## Data Availability

SPRINT clinical trial data is available through the NHLBI Biologic Specimen and Data Repository Information Coordinating Center

https://biolincc.nhlbi.nih.gov/studies/sprint

## Acknowledgements

We used Claude Opus 4.6 (Anthropic) in 2026 for editing of author-generated content to improve the clarity of the manuscript. The authors take responsibility for the integrity of the content generated.

## Disclaimer

The content is solely the responsibility of the authors and does not necessarily represent the official views of the National Institutes of Health.

