## Supplementary Online Content for "Effect of Intensive vs Standard Blood Pressure Control According to *APOE* ε4 Genotype: A Secondary Analysis of SPRINT"

Formatted for JAMA Neurology

### Supplemental Methods

#### Rationale for Addressing the Competing Risk of Death

In dementia research, the competing risk of death warrants particular attention because dementia typically manifests late in life when mortality rates are high. The impact of competing deaths on treatment effect estimation depends on two factors: the overall death rate during follow-up and how it compares with the event rate of the outcome of interest. In our study, during the primary trial follow-up (up to 5.1 years), the overall death rate was slightly higher than the dementia event rate but lower than the composite MCI or dementia event rate; thus, the impact of competing deaths was modest. During the extended follow-up, however, the death rate was 4- to 8-fold higher than the dementia event rate and 2- to 3-fold higher than the composite outcome event rate (eFigure 4). The impact of competing deaths is inherently larger for dementia than for the composite outcome, given the lower event rate of the former.

#### Limitations of Cause-Specific Hazard Ratios under Competing Risks

Cause-specific hazard ratios can produce misleading treatment effect estimates when competing deaths are frequent and differential between treatment groups. This is because the cause-specific approach censors deaths, effectively removing these individuals from the risk set. If one treatment group experiences higher mortality and individuals who died without dementia also had lower cognitive function, the remaining risk set in that group becomes smaller and potentially healthier, yielding fewer observed dementia events and an apparent cognitive benefit that is an artifact of differential mortality rather than a true protective effect. Furthermore, when the Cox model estimates a single HR that is influenced by the late follow-up period up to 11 years, where competing deaths dominated, these cause-specific estimates for dementia are not reliably interpretable. Also note that hazard ratio is not a causal estimand as it conditions on subjects being at risk at each follow-up time.

Despite these caveats, we report cause-specific hazard ratio as a primary estimand for two reasons. First, by adjusting for the risk set over time, it captures the etiological effect of treatment, which is often the effect of interest to clinicians. Second, the hazard ratio is widely used in medical journals, providing readers a familiar interpretation.

#### Competing Risk Framework Using the Aalen-Johansen Estimator

To address these limitations, we applied a competing risks framework using the Aalen-Johansen estimator to jointly estimate the cumulative incidence function (CIF) for both dementia and dementia-free death. These two CIFs should be interpreted jointly to assess whether a lower dementia risk in one treatment group may be explained by a higher competing risk of death, providing a more comprehensive depiction of treatment effects than cause-specific HRs alone. Particular caution is warranted when one group shows a lower risk of dementia but a higher risk of the composite outcome, or vice versa. In contrast to cause-specific hazard ratio, the risk difference and risk ratio derived from CIFs at time t are causal estimands, since the CIF is a marginal risk (unconditional on subjects remaining at risk).

#### *APOE* Genotyping Quality Control

*APOE* genotyping was performed at the Utah Genomics Core using TaqMan^®^ SNP Genotyping Assays (C____904973_10 for rs7412 and C___3084793_20 for rs429358). Of 9,361 SPRINT participants, 8,451 (90.3%) had stored DNA available. The overall genotyping call rate was 99.3%, with SNP-specific call rates of 99.4% for both rs7412 and rs429358 after rerunning initially failed samples. Concordance across three control samples was 100%. Of 8,451 samples assayed, 61 (0.7%) were excluded due to discordant or uninterpretable results.

#### Diplotype-to-ε4 Allele Coding

The ε2 allele is defined by the T allele at both rs429358 and rs7412; ε3 by the T allele at rs429358 and C allele at rs7412; and ε4 by the C allele at both rs429358 and rs7412.^26^ From these allele definitions, six diplotype combinations are possible: ε2/ε2, ε2/ε3, and ε3/ε3 were assigned an ε4 allele count of 0; ε3/ε4 and ε2/ε4 were assigned a count of 1; and ε4/ε4 was assigned a count of 2. Individuals heterozygous at both rs429358 and rs7412 (CT and CT) were classified as ε2/ε4 by standard convention and assigned an ε4 count of 1. Samples yielding discordant or uninterpretable genotype combinations at rs429358 and rs7412 were flagged for quality control review and excluded from analyses (N=61).

#### Hardy-Weinberg Equilibrium Assessment

Observed *APOE* genotype frequencies were tested for conformity with Hardy-Weinberg equilibrium (HWE) using chi-squared goodness-of-fit tests, overall and stratified by self-reported race/ethnicity. Overall HWE P = 0.35. Non-Hispanic White P = 0.9; Non-Hispanic Black P = 0.46; Hispanic P = 0.68. No significant deviation from HWE was observed, supporting the validity of the genotyping assay.

#### Proportional Hazards Assumption Assessment

For the primary analysis, no violation of the proportional hazards assumption was observed (Schoenfeld test: P = .91 for ε4 carriers; P = .09 for non-carriers).

#### Outcome Ascertainment

Cognitive function was assessed using a standardized battery that included the Montreal Cognitive Assessment (MoCA), the Logical Memory I and II subtests of the Wechsler Memory Scale, and the Digit Symbol Coding Test.^8^ Participants who screened positive for possible cognitive impairment based on predefined thresholds were referred for an extended evaluation, which included a comprehensive neuropsychological battery, functional assessment, and informant interview.^8, 9^

During the extended follow-up, cognitive status was ascertained by telephone with participants and their trusted informants, with the Modified Telephone Interview for Cognitive Status replacing the in-person MoCA as the screening instrument.^8^ Although some assessments differed between the in-person and telephone batteries, an expert adjudication panel of neuropsychologists and physicians, blinded to treatment assignment, applied the same standardized diagnostic criteria across the main trial and extended follow-up, reviewing all available data to classify participants as having no cognitive impairment, MCI, or dementia.^8^ The primary outcome was incident dementia. Secondary outcomes included MCI; the composite of MCI or dementia; the composite of MCI, dementia, or death; and all-cause mortality. MCI required two consecutive adjudicated classifications of MCI (eFigure 2).

### Supplemental Figures

#### eFigure 1. Timeline for follow-up and *APOE* ε4 genotyping in the Systolic Blood Pressure Intervention Trial.

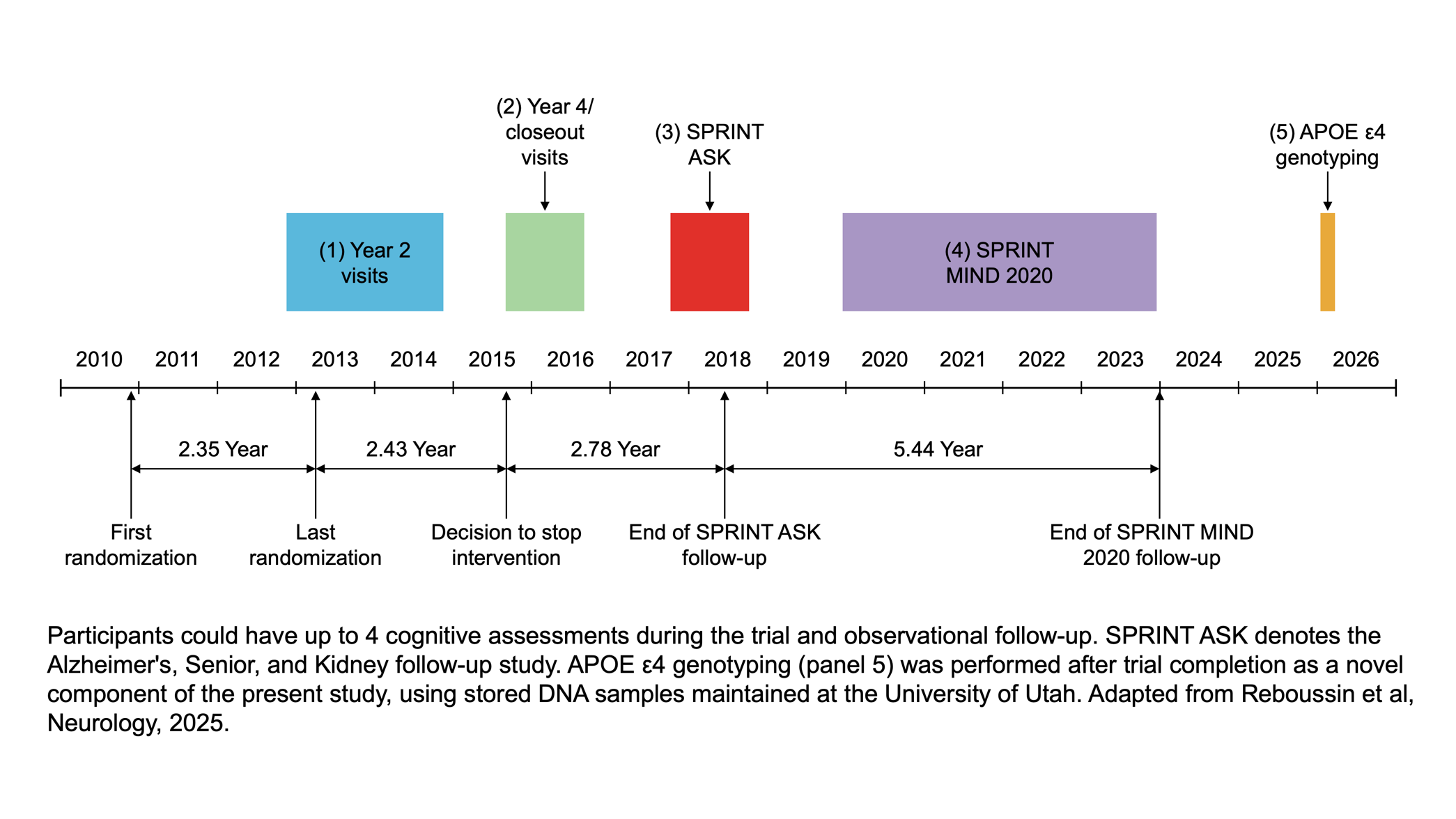

#### eFigure 2. Schematic depicting possible combinations of adjudication decisions including mild cognitive impairment (MCI).

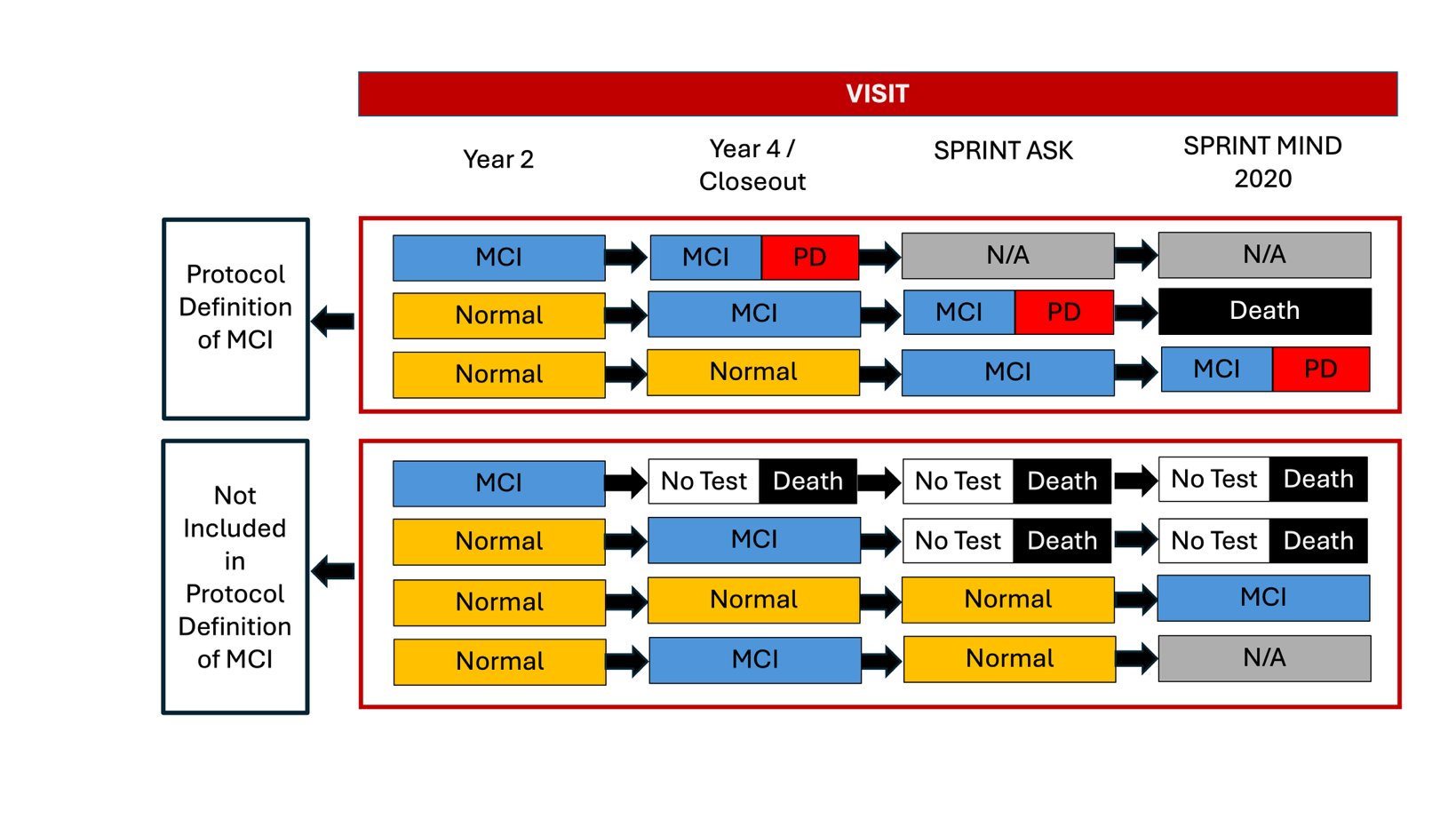

PD denotes dementia. Normal indicates no cognitive impairment. N/A indicates that the outcome definition of MCI is not dependent upon cognitive status at that particular visit. MCI required at least two consecutive adjudicated classifications of MCI (top panel). Adapted from Williamson et al, JAMA, 2019.

#### eFigure 3. Achieved Systolic Blood Pressure by *APOE* ε4 Status.

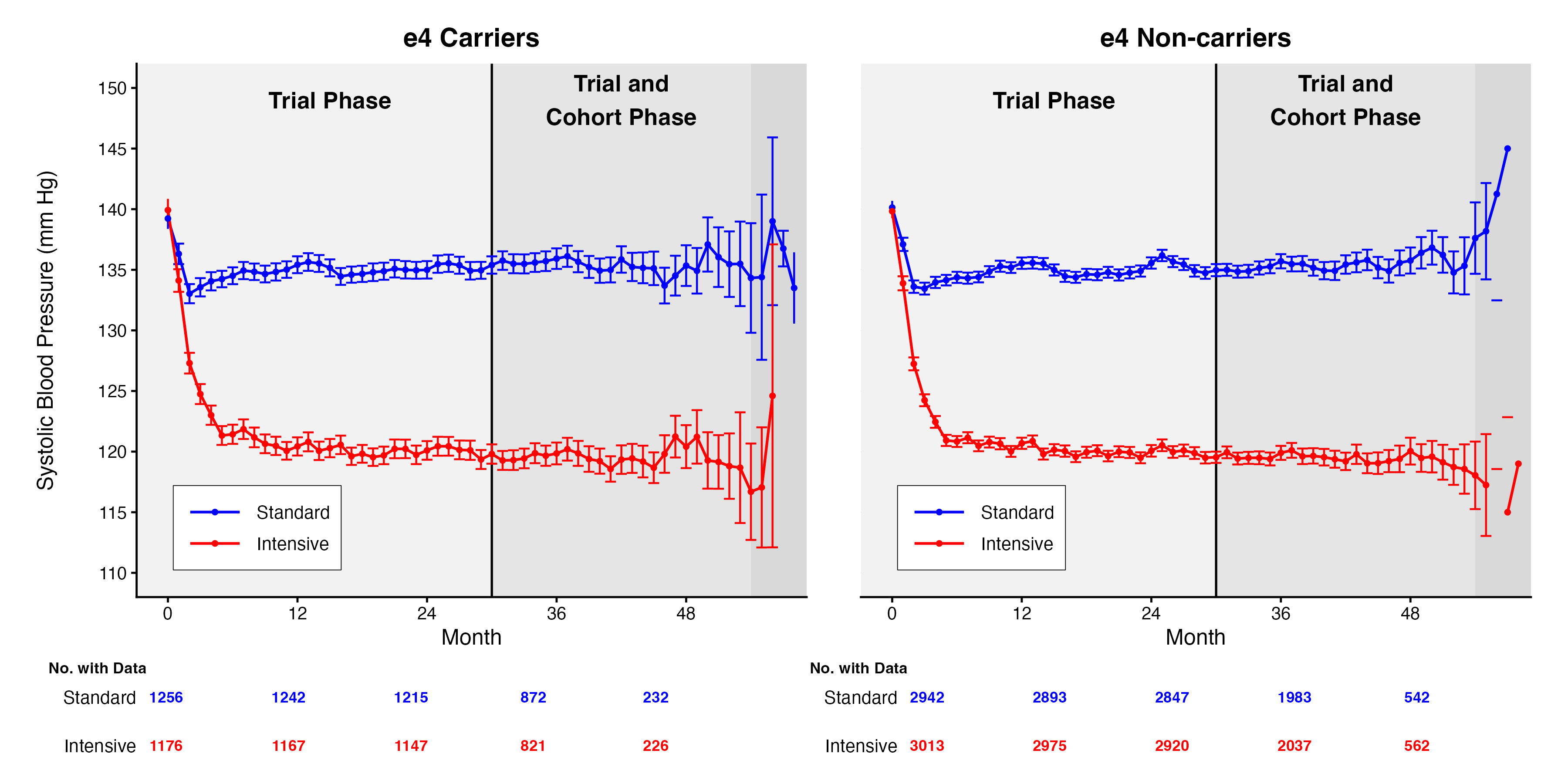

Mean achieved SBP over time by treatment group, stratified by *APOE* ε4 carrier status This figure demonstrates that the SBP separation between treatment groups was similar regardless of *APOE* status.

#### eFigure 4. Cumulative incidence curves by *APOE* ε4 carrier status for cognitive outcomes during extended follow-up.

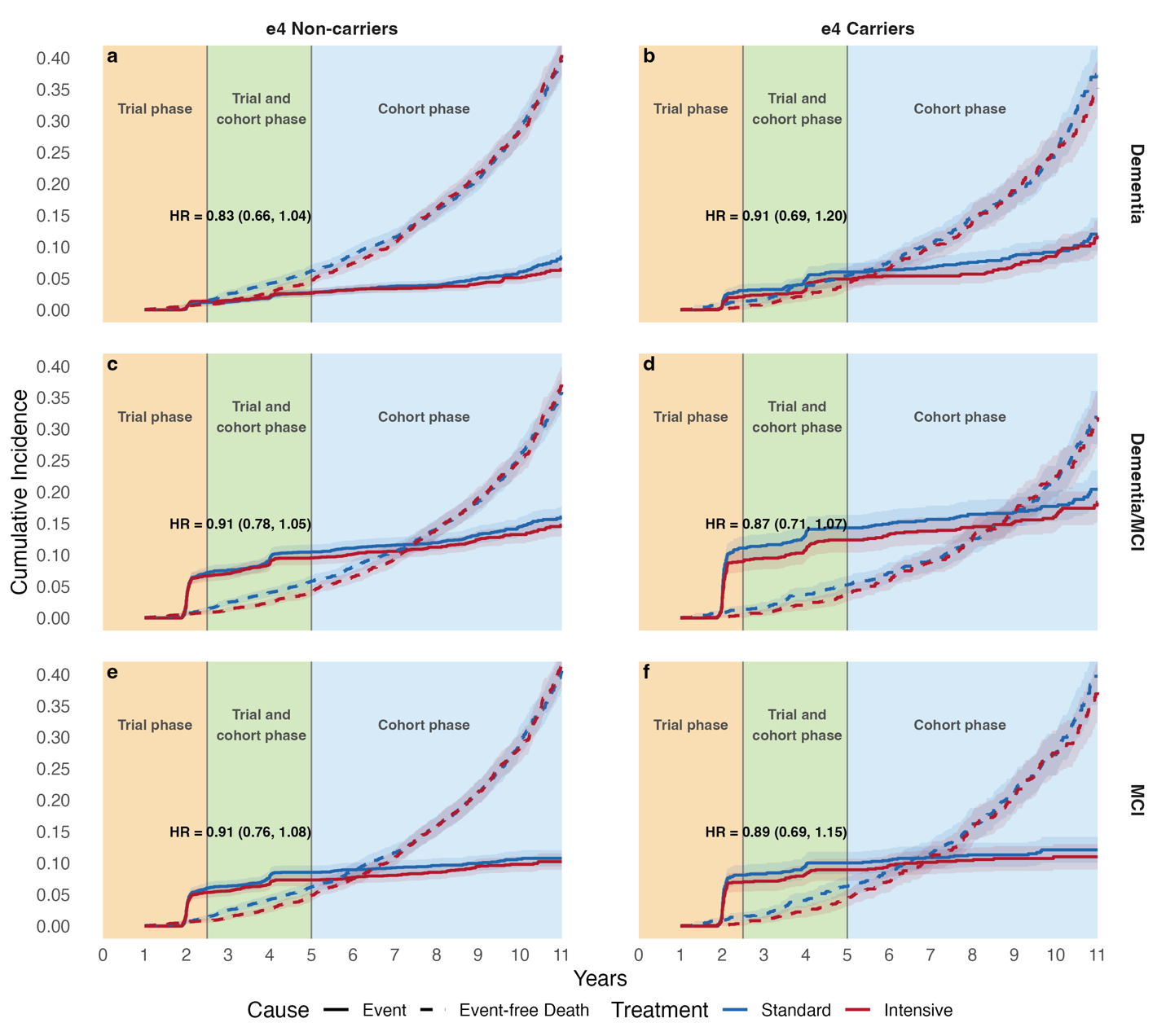

Cumulative incidence curves show the risk of event (e.g., dementia) and event-free death under the intensive vs standard SBP control for *APOE* ε4 carriers and non-carriers, estimated using a competing risk framework. Shaded areas indicate 95% confidence intervals.

#### eFigure 5. Risk Differences of Intensive vs Standard SBP Control on Cognitive Outcomes by *APOE* ε4 Carrier Status in the Primary Follow-up.

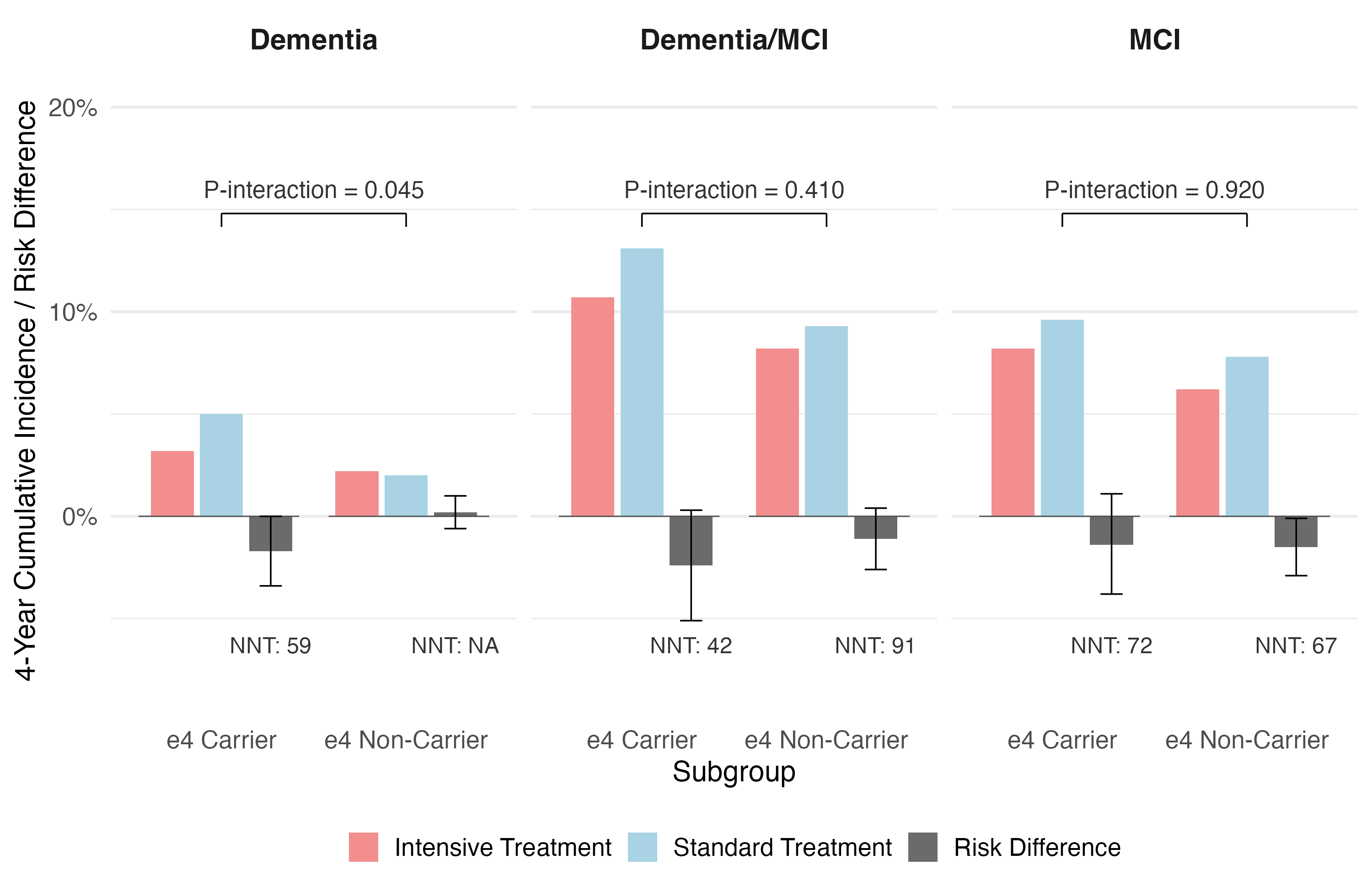

#### eFigure 6. Effects of Intensive vs Standard SBP Control on Cognitive Outcomes by *APOE* ε4 Carrier Status in the Extended Follow-up.

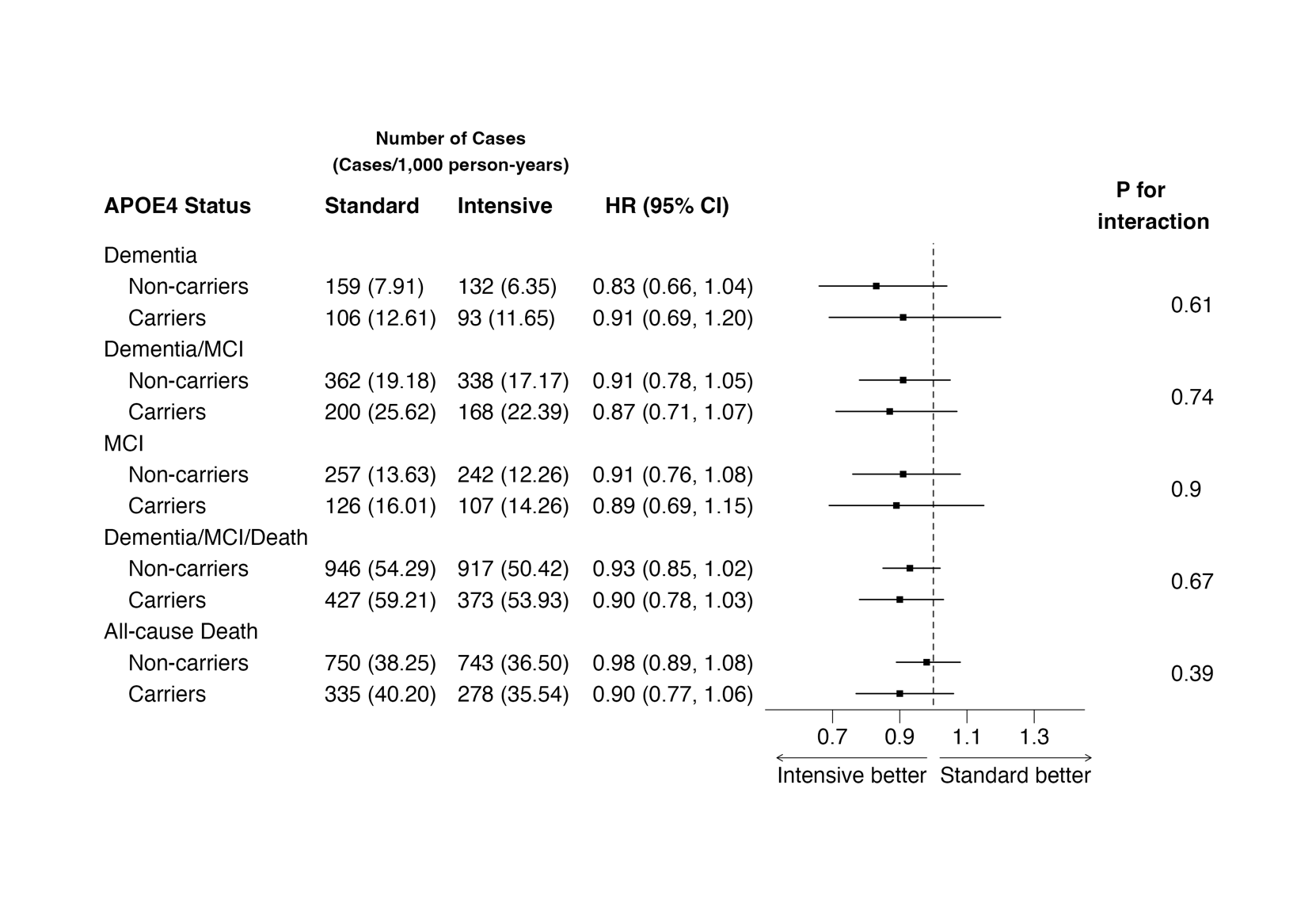

CI = confidence interval; HR = hazard ratio; MCI = mild cognitive impairment. Hazard ratios compare intensive SBP control (<120 mm Hg) with standard SBP control (<140 mm Hg) as referent. Interaction p-values are from Wald tests of the treatment × *APOE* ε4 interaction term. Event rates are per 1000 person-years (py); mortality rates are reported alongside clinical outcome rates to assess the impact of the competing event of death. These sensitivity analyses used the primary SPRINT follow-up and extended follow-up from SPRINT MIND 2020 without adjusting for baseline covariates. Death events prior to dementia or MCI are censored at the time of death.

### Supplemental Tables

#### eTable 1. Distribution of *APOE* Genotype by Race/Ethnicity.

|  | **Overall**  (N = 8,387) | **NH White**  (N = 4,943) | **NH Black**  (N = 2,384) | **Hispanic**  (N = 904) | **Other**  (N = 156) |
| --- | --- | --- | --- | --- | --- |
| **Panel A: Allele frequencies** | | | | | |
| ε2 | 0.08 | 0.08 | 0.11 | 0.05 | 0.07 |
| ε3 | 0.76 | 0.79 | 0.67 | 0.82 | 0.8 |
| ε4 | 0.16 | 0.13 | 0.22 | 0.13 | 0.13 |
| **Panel B: Observed genotype counts (expected under HWE)** | | | | | |
| ε2/ε2 | 67 (56.7) | 32 (28.4) | 34 (28.0) | 1 (2.3) | 0 (0.8) |
| ε2/ε3 | 1020 (1048.1) | 588 (592.9) | 342 (347.0) | 74 (74.2) | 16 (17.6) |
| ε2/ε4 | 225 (217.5) | 97 (99.4) | 107 (114.0) | 15 (12.2) | 6 (2.9) |
| ε3/ε3 | 4868 (4844.9) | 3097 (3096.8) | 1067 (1073.8) | 604 (601.7) | 100 (99.4) |
| ε3/ε4 | 1993 (2011.1) | 1043 (1038.5) | 724 (705.4) | 193 (197.4) | 33 (32.7) |
| ε4/ε4 | 214 (208.7) | 86 (87.1) | 110 (115.8) | 17 (16.2) | 1 (2.7) |
| **Panel C: HWE test** | | | | | |
| χ² (df = 3) | 3.29 | 0.59 | 2.6 | 1.53 | 5.33 |
| **P value** | **0.35** | **0.9** | **0.46** | **0.68** | **0.15** |

Allele frequencies are reported as proportions. Observed genotype counts are presented with expected counts under Hardy-Weinberg equilibrium in parentheses. Expected counts were calculated from observed allele frequencies assuming random mating: for alleles with frequencies p, q, and r (ε2, ε3, ε4), expected homozygote frequencies are p², q², r² and expected heterozygote frequencies are 2pq, 2pr, 2qr. The chi-squared goodness-of-fit test has 3 degrees of freedom for a 3-allele system (6 genotype classes minus 3 estimated allele frequencies). HWE testing was performed on the full genotyped sample (N = 8,390) rather than the analytic cohort (N = 7,733) to avoid selection bias from excluding participants with missing cognitive data. Three participants had missing race. NH = Non-Hispanic.

#### eTable 2. Baseline Characteristics of SPRINT and Current Study by *APOE* ε4 Genotype.

| **Characteristic** | **SPRINT**  N = 9,361^1^ | **Overall**  N = 7,733^1^ | **Non-carriers**  N = 5,492^1^ | **Heterozygote**  N = 2,049^1^ | **Homozygote**  N = 192^1^ | **Max ASMD**^2^ |
| --- | --- | --- | --- | --- | --- | --- |
| **Randomized Treatment** |  |  |  |  |  | 0.05 |
| Standard | 4,683 (50.0%) | 3,867 (50.0%) | 2,711 (49.4%) | 1,060 (51.7%) | 96 (50.0%) |  |
| Intensive | 4,678 (50.0%) | 3,866 (50.0%) | 2,781 (50.6%) | 989 (48.3%) | 96 (50.0%) |  |
| **Age** | 67.9 (9.4) | 68.0 (9.3) | 68.5 (9.4) | 66.9 (9.2) | 65.0 (8.3) | 0.40 |
| **Female** | 3,332 (35.6%) | 2,779 (35.9%) | 1,947 (35.5%) | 748 (36.5%) | 84 (43.8%) | 0.17 |
| **Race** |  |  |  |  |  | 0.54 |
| Hispanic | 984 (10.5%) | 820 (10.6%) | 618 (11.3%) | 187 (9.1%) | 15 (7.8%) |  |
| NH Black | 2,802 (29.9%) | 2,155 (27.9%) | 1,304 (23.7%) | 758 (37.0%) | 93 (48.4%) |  |
| NH White | 5,399 (57.7%) | 4,617 (59.7%) | 3,465 (63.1%) | 1,069 (52.2%) | 83 (43.2%) |  |
| Other | 176 (1.9%) | 141 (1.8%) | 105 (1.9%) | 35 (1.7%) | 1 (0.5%) |  |
| **Education Level** |  |  |  |  |  | 0.23 |
| College graduate | 3,636 (38.9%) | 3,136 (40.6%) | 2,305 (42.0%) | 770 (37.6%) | 61 (31.8%) |  |
| High school diploma or GED | 1,520 (16.3%) | 1,218 (15.8%) | 848 (15.4%) | 329 (16.1%) | 41 (21.4%) |  |
| Less than high school diploma | 876 (9.4%) | 688 (8.9%) | 482 (8.8%) | 190 (9.3%) | 16 (8.3%) |  |
| Some college (No degree) | 3,313 (35.5%) | 2,691 (34.8%) | 1,857 (33.8%) | 760 (37.1%) | 74 (38.5%) |  |
| **Private Insurance** | 3,980 (42.6%) | 3,416 (44.2%) | 2,449 (44.6%) | 893 (43.6%) | 74 (38.5%) | 0.12 |
| **Systolic Blood Pressure** | 139.7 (15.6) | 139.8 (15.6) | 139.9 (15.6) | 139.5 (15.7) | 139.0 (16.3) | 0.06 |
| **Diastolic Blood Pressure** | 78.1 (11.9) | 78.1 (11.9) | 77.8 (11.8) | 78.8 (12.0) | 79.1 (12.1) | 0.11 |
| **eGFR** | 71.8 (20.6) | 71.7 (20.4) | 71.3 (20.1) | 72.8 (21.0) | 73.0 (21.3) | 0.09 |
| **Serum Glucose** | 98.8 (13.5) | 98.8 (13.6) | 99.1 (13.5) | 98.2 (13.6) | 97.3 (13.7) | 0.13 |
| **Total Cholesterol** | 190.1 (41.2) | 190.0 (41.1) | 187.9 (40.7) | 194.4 (41.2) | 202.9 (43.6) | 0.35 |
| **HDL Cholesterol** | 52.9 (14.5) | 52.8 (14.4) | 53.1 (14.4) | 52.1 (14.3) | 51.8 (14.7) | 0.09 |
| **Statin Use** | 4,063 (43.4%) | 3,395 (43.9%) | 2,342 (42.6%) | 953 (46.5%) | 100 (52.1%) | 0.19 |
| **Aspirin Use** | 4,914 (52.5%) | 4,103 (53.1%) | 2,923 (53.2%) | 1,076 (52.5%) | 104 (54.2%) | 0.03 |
| **Smoking Status** |  |  |  |  |  | 0.26 |
| Current | 1,240 (13.3%) | 954 (12.3%) | 630 (11.5%) | 284 (13.9%) | 40 (20.8%) |  |
| Former | 3,973 (42.6%) | 3,308 (42.8%) | 2,382 (43.4%) | 846 (41.3%) | 80 (41.7%) |  |
| Never | 4,122 (44.2%) | 3,465 (44.8%) | 2,475 (45.1%) | 918 (44.8%) | 72 (37.5%) |  |
| **Body Mass Index** | 29.9 (5.8) | 29.8 (5.8) | 29.8 (5.7) | 29.9 (5.8) | 30.0 (6.8) | 0.03 |
| **ACEI or ARB** | 5,469 (58.4%) | 4,528 (58.6%) | 3,269 (59.5%) | 1,157 (56.5%) | 102 (53.1%) | 0.13 |
| **Beta Blocker** | 3,413 (36.5%) | 2,805 (36.3%) | 1,985 (36.1%) | 749 (36.6%) | 71 (37.0%) | 0.02 |
| **Loop Diuretic** | 546 (5.8%) | 448 (5.8%) | 306 (5.6%) | 130 (6.3%) | 12 (6.3%) | 0.03 |
| **Calcium Channel Blocker** | 3,319 (35.5%) | 2,680 (34.7%) | 1,906 (34.7%) | 712 (34.7%) | 62 (32.3%) | 0.05 |
| **Thiazide Diuretic** | 3,789 (40.5%) | 3,115 (40.3%) | 2,210 (40.2%) | 825 (40.3%) | 80 (41.7%) | 0.03 |
| **Alpha Blocker** | 940 (10.0%) | 775 (10.0%) | 575 (10.5%) | 186 (9.1%) | 14 (7.3%) | 0.11 |
| **Other Antihypertensive Medication** | 373 (4.0%) | 299 (3.9%) | 201 (3.7%) | 86 (4.2%) | 12 (6.3%) | 0.12 |
| **Family History of Heart Disease** |  |  |  |  |  | 0.15 |
| No | 3,093 (33.1%) | 2,510 (32.5%) | 1,742 (31.7%) | 706 (34.5%) | 62 (32.3%) |  |
| Unknown | 544 (5.8%) | 423 (5.5%) | 309 (5.6%) | 98 (4.8%) | 16 (8.3%) |  |
| Yes | 5,701 (61.1%) | 4,796 (62.1%) | 3,437 (62.6%) | 1,245 (60.8%) | 114 (59.4%) |  |
| **Framingham Risk Score** | 22.2 (15.3, 31.9) | 22.1 (15.3, 31.8) | 22.2 (15.3, 32.0) | 21.7 (15.1, 31.3) | 22.1 (14.7, 32.0) | 0.03 |
| **MoCA Score** | 23.0 (20.0, 26.0) | 24.0 (21.0, 26.0) | 24.0 (21.0, 26.0) | 23.0 (21.0, 26.0) | 23.0 (19.0, 26.0) | 0.13 |
| **Digit Symbol Coding Test** | 50.8 (15.3) | 51.4 (15.3) | 51.6 (15.3) | 51.2 (15.1) | 49.1 (15.1) | 0.16 |
| **Logical Memory Delayed Recall (LM2)** | 8.2 (3.4) | 8.3 (3.3) | 8.4 (3.3) | 8.2 (3.4) | 7.7 (3.7) | 0.20 |
| ^1^n (%); Mean (SD); Median (Q1, Q3)  ^2^The maximum of 3 pairwise ASMD among ε4 non-carriers, heterozygotes, and homozygotes. | | | | | | |

#### eTable 3. Assessing Effect Heterogeneity of Intensive vs Standard SBP Control by *APOE* Genotype.

| **Outcome** | **APOE^2^** | **Intensive Arm** | | | **Standard Arm** | | | **Intensive vs. Standard** | | | | **Interaction P-value** |
| --- | --- | --- | --- | --- | --- | --- | --- | --- | --- | --- | --- | --- |
|  |  | **N** | **Events** | **^1^Cases/**  **1,000 pys** | **N** | **Events** | **Cases/**  **1,000 pys** | **HR** | | **Lower**  **95% CL** | **Upper**  **95% CL** |  |
| Dementia | ε2 carriers | 538 | 6 | 2.28 | 461 | 16 | 6.94 | 0.35 | 0.14 | | 0.89 | 0.06 |
|  | ε3 carriers | 2243 | 76 | 6.95 | 2250 | 72 | 6.63 | 1.05 | 0.76 | | 1.45 |  |
|  | ε4 carriers | 1085 | 51 | 9.7 | 1156 | 72 | 13.18 | 0.73 | 0.51 | | 1.04 |  |
| MCI/Dementia | ε2 carriers | 538 | 39 | 15.34 | 461 | 61 | 28.1 | 0.55 | 0.36 | | 0.82 | 0.03 |
|  | ε3 carriers | 2243 | 203 | 19.21 | 2250 | 204 | 19.53 | 0.99 | 0.82 | | 1.21 |  |
|  | ε4 carriers | 1085 | 124 | 24.57 | 1156 | 157 | 30.22 | 0.82 | 0.65 | | 1.04 |  |
| MCI | ε2 carriers | 538 | 35 | 13.78 | 461 | 50 | 23.12 | 0.59 | 0.39 | | 0.91 | 0.24 |
|  | ε3 carriers | 2243 | 143 | 13.59 | 2250 | 158 | 15.19 | 0.9 | 0.72 | | 1.13 |  |
|  | ε4 carriers | 1085 | 87 | 17.36 | 1156 | 105 | 20.42 | 0.86 | 0.65 | | 1.15 |  |
| MCI/Dementia/ death | ε2 carriers | 538 | 66 | 32.98 | 461 | 84 | 49.17 | 0.67 | 0.49 | | 0.93 | 0.16 |
|  | ε3 carriers | 2243 | 313 | 37.63 | 2250 | 348 | 42.01 | 0.91 | 0.78 | | 1.06 |  |
|  | ε4 carriers | 1085 | 172 | 42.97 | 1156 | 218 | 52.36 | 0.77 | 0.63 | | 0.94 |  |
| All-cause death | ε2 carriers | 538 | 31 | 14.61 | 461 | 28 | 15.18 | 1.08 | 0.65 | | 1.8 | 0.38 |
|  | ε3 carriers | 2243 | 137 | 15.38 | 2250 | 167 | 18.83 | 0.89 | 0.71 | | 1.12 |  |
|  | ε4 carriers | 1085 | 62 | 14.24 | 1156 | 79 | 17.29 | 0.72 | 0.51 | | 1.01 |  |
| ^1^Numbers are counts and event rates (number of cases per 1,000 person-years [pys]). The primary outcome was the first occurrence of dementia. MCI = mild cognitive impairment, CL = Confidence limit. | | | | | | | | | | | | |
| ^2^ε2 carriers = ε2/ε2 or ε3/ε2; ε3 carriers = ε3/ε3; ε4 carriers = ε4/ε2 or ε4/ε4 or ε4/ε3. | | | | | | | | | | | | |

#### eTable 4. Association between *APOE* ε4 Carrier and Incident MCI and Dementia Outcomes.

| **Outcome** | **Characteristic** | **Model 1**  **Unadjusted HR**  **(95% CI)** | **Model 2 Demographic-Adjusted HR**  **(95% CI)** |
| --- | --- | --- | --- |
| **Dementia** | **PRIMARY EXPOSURE** | | |
|  | ***APOE* ε4 Carrier** | 1.82 (1.44, 2.29) | 2.22 (1.76, 2.81) |
|  | **DEMOGRAPHIC COVARIATES** | | |
|  | **Age** | — | 1.13 (1.12, 1.15) |
|  | **Sex - Female** | — | 1.02 (0.81, 1.29) |
|  | **Race/Ethnicity** | | |
|  | Non-Hispanic Black | — | 1.48 (1.11, 1.98) |
|  | Hispanic | — | 2.86 (2.07, 3.94) |
|  | Other | — | 1.09 (0.4, 2.93) |
| **MCI or Dementia** | **PRIMARY EXPOSURE** | | |
|  | ***APOE* ε4 Carrier** | 1.39 (1.2, 1.61) | 1.54 (1.33, 1.79) |
|  | **DEMOGRAPHIC COVARIATES** | | |
|  | **Age** | — | 1.1 (1.09, 1.11) |
|  | **Sex - Female** | — | 0.85 (0.73, 0.98) |
|  | **Race/Ethnicity** | | |
|  | Non-Hispanic Black | — | 2.18 (1.84, 2.57) |
|  | Hispanic | — | 2.7 (2.19, 3.33) |
|  | Other | — | 1.79 (1.1, 2.91) |
| **MCI** | **PRIMARY EXPOSURE** | | |
|  | ***APOE* ε4 Carrier** | 1.25 (1.05, 1.48) | 1.34 (1.13, 1.6) |
|  | **DEMOGRAPHIC COVARIATES** | | |
|  | **Age** | — | 1.09 (1.08, 1.1) |
|  | **Sex - Female** | — | 0.81 (0.69, 0.97) |
|  | **Race/Ethnicity** | | |
|  | Non-Hispanic Black | — | 2.46 (2.03, 2.97) |
|  | Hispanic | — | 2.74 (2.14, 3.53) |
|  | Other | — | 1.92 (1.1, 3.34) |

#### eTable 5. Association Between *APOE* ε4 Carrier and Incident MCI or Dementia or death outcome and all-cause mortality.

| **Outcome** | **Characteristic** | **Model 1**  **Unadjusted HR**  **(95% CI)** | **Model 2 Demographic-Adjusted HR**  **(95% CI)** |
| --- | --- | --- | --- |
| **MCI or Dementia or Death** | **PRIMARY EXPOSURE** | | |
|  | ***APOE* ε4 Carrier** | 1.18 (1.06, 1.32) | 1.3 (1.16, 1.46) |
|  | **DEMOGRAPHIC COVARIATES** | | |
|  | **Age** | — | 1.09 (1.08, 1.1) |
|  | **Sex - Female** | — | 0.72 (0.64, 0.81) |
|  | **Race/Ethnicity** | | |
|  | Non-Hispanic Black | — | 1.88 (1.65, 2.13) |
|  | Hispanic | — | 1.77 (1.49, 2.11) |
|  | Other | — | 1.41 (0.94, 2.12) |
| **All-cause Death** | **PRIMARY EXPOSURE** | | |
|  | ***APOE* ε4 Carrier** | 0.94 (0.8, 1.11) | 1.03 (0.87, 1.22) |
|  | **DEMOGRAPHIC COVARIATES** | | |
|  | **Age** | — | 1.08 (1.07, 1.09) |
|  | **Sex - Female** | — | 0.6 (0.51, 0.71) |
|  | **Race/Ethnicity** | | |
|  | Non-Hispanic Black | — | 1.33 (1.11, 1.59) |
|  | Hispanic | — | 0.8 (0.59, 1.08) |
|  | Other | — | 0.97 (0.5, 1.89) |

#### eTable 6. Association Between *APOE* ε4 Count and Incident MCI or Dementia Outcomes.

| **Outcome** | **Characteristic** | **Model 1**  **Unadjusted HR**  **(95% CI)** | **Model 2 Demographic-Adjusted HR**  **(95% CI)** |
| --- | --- | --- | --- |
| **Dementia** | **PRIMARY EXPOSURE** | | |
|  | ***APOE* ε4 Count** | 1.66 (1.37, 2) | 2.03 (1.67, 2.46) |
|  | **DEMOGRAPHIC COVARIATES** | | |
|  | **Age** | — | 1.13 (1.12, 1.15) |
|  | **Sex - Female** | — | 1.01 (0.8, 1.28) |
|  | **Race/Ethnicity** | | |
|  | Non-Hispanic Black | — | 1.48 (1.11, 1.97) |
|  | Hispanic | — | 2.85 (2.07, 3.93) |
|  | Other | — | 1.1 (0.41, 2.97) |
| **MCI or Dementia** | **PRIMARY EXPOSURE** | | |
|  | ***APOE* ε4 Count** | 1.34 (1.18, 1.51) | 1.49 (1.31, 1.69) |
|  | **DEMOGRAPHIC COVARIATES** | | |
|  | **Age** | — | 1.1 (1.09, 1.11) |
|  | **Sex - Female** | — | 0.85 (0.73, 0.98) |
|  | **Race/Ethnicity** | | |
|  | Non-Hispanic Black | — | 2.17 (1.84, 2.56) |
|  | Hispanic | — | 2.7 (2.19, 3.33) |
|  | Other | — | 1.8 (1.11, 2.93) |
| **MCI** | **PRIMARY EXPOSURE** | | |
|  | ***APOE* ε4 Count** | 1.23 (1.06, 1.42) | 1.32 (1.14, 1.54) |
|  | **DEMOGRAPHIC COVARIATES** | | |
|  | **Age** | — | 1.09 (1.08, 1.1) |
|  | **Sex - Female** | — | 0.81 (0.68, 0.97) |
|  | **Race/Ethnicity** | | |
|  | Non-Hispanic Black | — | 2.45 (2.02, 2.96) |
|  | Hispanic | — | 2.75 (2.14, 3.53) |
|  | Other | — | 1.92 (1.1, 3.35) |

#### eTable 7. Association Between *APOE* ε4 Count and Incident MCI or Dementia or death outcome and all-cause mortality.

| **Outcome** | **Characteristic** | **Model 1**  **Unadjusted HR**  **(95% CI)** | **Model 2 Demographic-Adjusted HR**  **(95% CI)** |
| --- | --- | --- | --- |
| **MCI or Dementia or Death** | **PRIMARY EXPOSURE** | | |
|  | ***APOE* ε4 Count** | 1.14 (1.04, 1.26) | 1.26 (1.14, 1.39) |
|  | **DEMOGRAPHIC COVARIATES** | | |
|  | **Age** | — | 1.09 (1.08, 1.1) |
|  | **Sex - Female** | — | 0.72 (0.64, 0.81) |
|  | **Race/Ethnicity** | | |
|  | Non-Hispanic Black | — | 1.87 (1.65, 2.13) |
|  | Hispanic | — | 1.77 (1.49, 2.11) |
|  | Other | — | 1.42 (0.94, 2.13) |
| **All-cause Death** | **PRIMARY EXPOSURE** | | |
|  | ***APOE* ε4 Count** | 0.93 (0.8, 1.07) | 1.01 (0.87, 1.18) |
|  | **DEMOGRAPHIC COVARIATES** | | |
|  | **Age** | — | 1.08 (1.07, 1.08) |
|  | **Sex - Female** | — | 0.6 (0.51, 0.71) |
|  | **Race/Ethnicity** | | |
|  | Non-Hispanic Black | — | 1.33 (1.11, 1.59) |
|  | Hispanic | — | 0.8 (0.59, 1.08) |
|  | Other | — | 0.98 (0.5, 1.89) |

#### eTable 8. Association Between *APOE* ε2 Carrier and Incident MCI or Dementia Outcomes.

| **Outcome** | **Characteristic** | **Model 1**  **Unadjusted HR**  **(95% CI)** | **Model 2 Demographic-Adjusted HR**  **(95% CI)** |
| --- | --- | --- | --- |
| **Dementia** | **PRIMARY EXPOSURE** | | |
|  | ***APOE* ε2 Carrier** | 0.66 (0.46, 0.95) | 0.62 (0.43, 0.90) |
|  | **DEMOGRAPHIC COVARIATES** | | |
|  | **Age** | — | 1.13 (1.11, 1.14) |
|  | **Sex - Female** | — | 1.01 (0.80, 1.28) |
|  | **Race/Ethnicity** | | |
|  | Non-Hispanic Black | — | 1.73 (1.30, 2.30) |
|  | Hispanic | — | 2.82 (2.05, 3.90) |
|  | Other | — | 1.10 (0.41, 2.96) |
| **MCI or Dementia** | **PRIMARY EXPOSURE** | | |
|  | ***APOE* ε2 Carrier** | 0.94 (0.77, 1.14) | 0.89 (0.73, 1.08) |
|  | **DEMOGRAPHIC COVARIATES** | | |
|  | **Age** | — | 1.10 (1.09, 1.11) |
|  | **Sex - Female** | — | 0.85 (0.73, 0.98) |
|  | **Race/Ethnicity** | | |
|  | Non-Hispanic Black | — | 2.33 (1.98, 2.75) |
|  | Hispanic | — | 2.68 (2.17, 3.30) |
|  | Other | — | 1.81 (1.12, 2.95) |
| **MCI** | **PRIMARY EXPOSURE** | | |
|  | ***APOE* ε2 Count** | 1.05 (0.84, 1.30) | 0.98 (0.79, 1.23) |
|  | **DEMOGRAPHIC COVARIATES** | | |
|  | **Age** | — | 1.09 (1.08, 1.10) |
|  | **Sex - Female** | — | 0.81 (0.68, 0.96) |
|  | **Race/Ethnicity** | | |
|  | Non-Hispanic Black | — | 2.56 (2.12, 3.10) |
|  | Hispanic | — | 2.73 (2.12, 3.50) |
|  | Other | — | 1.92 (1.10, 3.35) |

#### eTable 9. Association Between *APOE* ε2 Carrier and Incident MCI or Dementia or Death and All-cause Mortality.

| **Outcome** | **Characteristic** | **Model 1**  **Unadjusted HR**  **(95% CI)** | **Model 2 Demographic-Adjusted HR**  **(95% CI)** |
| --- | --- | --- | --- |
| **MCI or Dementia or Death** | **PRIMARY EXPOSURE** | | |
|  | ***APOE* ε2 Carrier** | 0.93 (0.8, 1.08) | 0.9 (0.77, 1.04) |
|  | **DEMOGRAPHIC COVARIATES** | | |
|  | **Age** | — | 1.09 (1.08, 1.1) |
|  | **Sex - Female** | — | 0.72 (0.64, 0.81) |
|  | **Race/Ethnicity** | | |
|  | Non-Hispanic Black | — | 1.96 (1.73, 2.22) |
|  | Hispanic | — | 1.75 (1.47, 2.09) |
|  | Other | — | 1.43 (0.95, 2.15) |
| **All-cause Death** | **PRIMARY EXPOSURE** | | |
|  | ***APOE* ε2 Carrier** | 0.93 (0.75, 1.16) | 0.92 (0.74, 1.15) |
|  | **DEMOGRAPHIC COVARIATES** | | |
|  | **Age** | — | 1.08 (1.07, 1.08) |
|  | **Sex - Female** | — | 0.6 (0.51, 0.71) |
|  | **Race/Ethnicity** | | |
|  | Non-Hispanic Black | — | 1.34 (1.11, 1.6) |
|  | Hispanic | — | 0.79 (0.58, 1.07) |
|  | Other | — | 0.98 (0.5, 1.89) |

#### eTable 10. Association Between *APOE* ε2 Count and Incident MCI or Dementia Outcomes.

| **Outcome** | **Characteristic** | **Model 1**  **Unadjusted HR**  **(95% CI)** | **Model 2 Demographic-Adjusted HR**  **(95% CI)** |
| --- | --- | --- | --- |
| **Dementia** | **PRIMARY EXPOSURE** | | |
|  | ***APOE* ε2 Count** | 0.65 (0.46, 0.92) | 0.61 (0.43, 0.87) |
|  | **DEMOGRAPHIC COVARIATES** | | |
|  | **Age** | — | 1.13 (1.11, 1.14) |
|  | **Sex - Female** | — | 1.01 (0.8, 1.28) |
|  | **Race/Ethnicity** | | |
|  | Non-Hispanic Black | — | 1.73 (1.3, 2.31) |
|  | Hispanic | — | 2.82 (2.04, 3.89) |
|  | Other | — | 1.1 (0.41, 2.96) |
| **MCI or Dementia** | **PRIMARY EXPOSURE** | | |
|  | ***APOE* ε2 Count** | 0.93 (0.78, 1.12) | 0.88 (0.74, 1.06) |
|  | **DEMOGRAPHIC COVARIATES** | | |
|  | **Age** | — | 1.1 (1.09, 1.11) |
|  | **Sex - Female** | — | 0.85 (0.73, 0.98) |
|  | **Race/Ethnicity** | | |
|  | Non-Hispanic Black | — | 2.34 (1.98, 2.75) |
|  | Hispanic | — | 2.68 (2.17, 3.3) |
|  | Other | — | 1.81 (1.11, 2.95) |
| **MCI** | **PRIMARY EXPOSURE** | | |
|  | ***APOE* ε2 Count** | 1.04 (0.85, 1.28) | 0.98 (0.8, 1.2) |
|  | **DEMOGRAPHIC COVARIATES** | | |
|  | **Age** | — | 1.09 (1.08, 1.1) |
|  | **Sex - Female** | — | 0.81 (0.68, 0.96) |
|  | **Race/Ethnicity** | | |
|  | Non-Hispanic Black | — | 2.56 (2.12, 3.1) |
|  | Hispanic | — | 2.72 (2.12, 3.5) |
|  | Other | — | 1.92 (1.1, 3.35) |

#### eTable 11. Association Between *APOE* ε2 Count and Incident MCI or Dementia or Death and All-cause Mortality.

| **Outcome** | **Characteristic** | **Model 1**  **Unadjusted HR**  **(95% CI)** | **Model 2 Demographic-Adjusted HR**  **(95% CI)** |
| --- | --- | --- | --- |
| **MCI or Dementia or Death** | **PRIMARY EXPOSURE** | | |
|  | ***APOE* ε2 Count** | 0.95 (0.82, 1.09) | 0.91 (0.79, 1.04) |
|  | **DEMOGRAPHIC COVARIATES** | | |
|  | **Age** | — | 1.09 (1.08, 1.1) |
|  | **Sex - Female** | — | 0.72 (0.64, 0.81) |
|  | **Race/Ethnicity** | | |
|  | Non-Hispanic Black | — | 1.96 (1.73, 2.22) |
|  | Hispanic | — | 1.75 (1.47, 2.09) |
|  | Other | — | 1.43 (0.95, 2.14) |
| **All-cause Death** | **PRIMARY EXPOSURE** | | |
|  | ***APOE* ε2 Count** | 0.96 (0.78, 1.17) | 0.94 (0.77, 1.15) |
|  | **DEMOGRAPHIC COVARIATES** | | |
|  | **Age** | — | 1.08 (1.07, 1.08) |
|  | **Sex - Female** | — | 0.6 (0.51, 0.71) |
|  | **Race/Ethnicity** | | |
|  | Non-Hispanic Black | — | 1.33 (1.11, 1.6) |
|  | Hispanic | — | 0.79 (0.59, 1.07) |
|  | Other | — | 0.98 (0.5, 1.89) |

#### eTable 12. Assessing effect heterogeneity of intensive vs standard SBP control by ε4 carrier status during the extended follow-up using a competing risk framework.

| Time | Outcome | ε4 non-carriers | | | | ε4 Carriers | | | |
| --- | --- | --- | --- | --- | --- | --- | --- | --- | --- |
|  |  | Risk | | RD^1^  (95% CI) | RR  (95% CI) | Risk | | RD  (95% CI) | RR  (95% CI) |
|  |  | Standard (N=2716) | Intensive (N=2782) |  |  | Standard (N=1157) | Intensive (N=1086) |  |  |
| **Primary Outcome** | | | | | | | | | |
| 4 years^2^ | PD | 0.019 | 0.021 | 0.002 (-0.006, 0.009) | 1.09 (0.75, 1.59) | 0.048 | 0.031 | -0.016 (-0.033, 0.000) | 0.66 (0.43, 1.01) |
|  | PD-free death | 0.042 | 0.028 | -0.014 (-0.024, -0.004) | 0.68 (0.51, 0.90) | 0.038 | 0.022 | -0.016 (-0.030, -0.001) | 0.59 (0.36, 0.97) |
| 6 years | PD | 0.032 | 0.032 | -0.000 (-0.010, 0.009) | 0.99 (0.73, 1.34) | 0.064 | 0.054 | -0.010 (-0.030, 0.011) | 0.85 (0.60, 1.20) |
|  | PD-free death | 0.088 | 0.074 | -0.014 (-0.029, 0.002) | 0.84 (0.70, 1.02) | 0.076 | 0.066 | -0.010 (-0.033, 0.013) | 0.87 (0.62, 1.20) |
| **Secondary Outcomes** | | | | | | | | | |
| 4 years | PD or MCI | 0.095 | 0.086 | -0.009 (-0.024, 0.007) | 0.91 (0.77, 1.08) | 0.133 | 0.108 | -0.024 (-0.052, 0.003) | 0.82 (0.65, 1.02) |
|  | PD or MCI-free death | 0.04 | 0.025 | -0.015 (-0.025, -0.006) | 0.63 (0.46, 0.85) | 0.037 | 0.02 | -0.017 (-0.031, -0.003) | 0.55 (0.33, 0.92) |
| 6 years | PD or MCI | 0.111 | 0.101 | -0.009 (-0.026, 0.007) | 0.92 (0.78, 1.07) | 0.15 | 0.133 | -0.018 (-0.047, 0.012) | 0.88 (0.71, 1.09) |
|  | PD or MCI-free death | 0.081 | 0.066 | -0.016 (-0.031, -0.001) | 0.81 (0.66, 0.99) | 0.072 | 0.059 | -0.013 (-0.036, 0.009) | 0.82 (0.58, 1.15) |
| 4 years | MCI | 0.08 | 0.067 | -0.013 (-0.027, 0.001) | 0.84 (0.69, 1.02) | 0.096 | 0.083 | -0.014 (-0.038, 0.010) | 0.86 (0.66, 1.12) |
|  | MCI-free death | 0.042 | 0.028 | -0.014 (-0.024, -0.004) | 0.67 (0.50, 0.89) | 0.041 | 0.022 | -0.018 (-0.033, -0.004) | 0.55 (0.33, 0.90) |
| 6 years | MCI | 0.089 | 0.078 | -0.012 (-0.027, 0.003) | 0.87 (0.72, 1.04) | 0.105 | 0.096 | -0.009 (-0.035, 0.016) | 0.91 (0.71, 1.18) |
|  | MCI-free death | 0.088 | 0.075 | -0.013 (-0.028, 0.003) | 0.86 (0.71, 1.04) | 0.089 | 0.07 | -0.019 (-0.043, 0.006) | 0.79 (0.58, 1.08) |

^1^CI = confidence interval; RD = risk difference; RR = risk ratio; MCI = mild cognitive impairment. RRs and RDs compare intensive SBP control (<120 mm Hg) with standard SBP control (<140 mm Hg) as referent.

^2^Estimates at 4 years from the extended follow-up data are very close to but not the same as those from primary follow-up data since 735 participants were censored before 4 years in the primary follow-up data but after 4 years in the extended follow-up data**.**

#### eTable 13. Association Between *APOE* ε4 Carrier and Incident MCI or Dementia Outcomes During the Extended Follow-up.

| **Outcome** | **Characteristic** | **Model 1**  **Unadjusted HR**  **(95% CI)** | **Model 2 Demographic-Adjusted HR**  **(95% CI)** |
| --- | --- | --- | --- |
| **Dementia** | **PRIMARY EXPOSURE** | | |
|  | ***APOE* ε4 Carrier** | 1.7 (1.42, 2.03) | 2.18 (1.82, 2.62) |
|  | **DEMOGRAPHIC COVARIATES** | | |
|  | **Age** | — | 1.12 (1.11, 1.13) |
|  | **Sex - Female** | — | 1.1 (0.92, 1.32) |
|  | **Race/Ethnicity** | | |
|  | Non-Hispanic Black | — | 1.17 (0.93, 1.47) |
|  | Hispanic | — | 2.14 (1.64, 2.78) |
|  | Other | — | 0.94 (0.42, 2.11) |
| **MCI or Dementia** | **PRIMARY EXPOSURE** | | |
|  | ***APOE* ε4 Carrier** | 1.31 (1.16, 1.49) | 1.5 (1.32, 1.7) |
|  | **DEMOGRAPHIC COVARIATES** | | |
|  | **Age** | — | 1.1 (1.09, 1.1) |
|  | **Sex - Female** | — | 0.84 (0.74, 0.96) |
|  | **Race/Ethnicity** | | |
|  | Non-Hispanic Black | — | 1.97 (1.71, 2.28) |
|  | Hispanic | — | 2.31 (1.92, 2.78) |
|  | Other | — | 1.44 (0.91, 2.27) |
| **MCI** | **PRIMARY EXPOSURE** | | |
|  | ***APOE* ε4 Carrier** | 1.17 (1, 1.36) | 1.24 (1.06, 1.45) |
|  | **DEMOGRAPHIC COVARIATES** | | |
|  | **Age** | — | 1.08 (1.07, 1.09) |
|  | **Sex - Female** | — | 0.78 (0.67, 0.91) |
|  | **Race/Ethnicity** | | |
|  | Non-Hispanic Black | — | 2.36 (2.00, 2.80) |
|  | Hispanic | — | 2.36 (1.88, 2.97) |
|  | Other | — | 1.56 (0.92, 2.67) |

#### eTable 14. Association Between *APOE* ε4 Carrier and Incident MCI or Dementia or Death and All-cause Mortality During the Extended Follow-up.

| **Outcome** | **Characteristic** | **Model 1**  **Unadjusted HR**  **(95% CI)** | **Model 2 Demographic-Adjusted HR**  **(95% CI)** |
| --- | --- | --- | --- |
| **MCI or Dementia or Death** | **PRIMARY EXPOSURE** | | |
|  | ***APOE* ε4 Carrier** | 1.07 (0.98, 1.16) | 1.24 (1.14, 1.35) |
|  | **DEMOGRAPHIC COVARIATES** | | |
|  | **Age** | — | 1.09 (1.08, 1.09) |
|  | **Sex - Female** | — | 0.75 (0.69, 0.82) |
|  | **Race/Ethnicity** | | |
|  | Non-Hispanic Black | — | 1.58 (1.44, 1.73) |
|  | Hispanic | — | 1.31 (1.14, 1.5) |
|  | Other | — | 1.06 (0.77, 1.46) |
| **All-cause Death** | **PRIMARY EXPOSURE** | | |
|  | ***APOE* ε4 Carrier** | 1 (0.91, 1.1) | 1.16 (1.06, 1.28) |
|  | **DEMOGRAPHIC COVARIATES** | | |
|  | **Age** | — | 1.09 (1.08, 1.09) |
|  | **Sex - Female** | — | 0.71 (0.65, 0.78) |
|  | **Race/Ethnicity** | | |
|  | Non-Hispanic Black | — | 1.32 (1.19, 1.47) |
|  | Hispanic | — | 0.89 (0.74, 1.06) |
|  | Other | — | 0.87 (0.59, 1.29) |

#### eTable 15. Association Between *APOE* ε2 Carrier and Incident MCI or Dementia Outcomes During the Extended Follow-up.

| **Outcome** | **Characteristic** | **Model 1**  **Unadjusted HR**  **(95% CI)** | **Model 2 Demographic-Adjusted HR**  **(95% CI)** |
| --- | --- | --- | --- |
| **Dementia** | **PRIMARY EXPOSURE** | | |
|  | ***APOE* ε2 Carrier** | 0.86 (0.67, 1.11) | 0.85 (0.66, 1.1) |
|  | **DEMOGRAPHIC COVARIATES** | | |
|  | **Age** | — | 1.12 (1.11, 1.13) |
|  | **Sex - Female** | — | 1.1 (0.91, 1.32) |
|  | **Race/Ethnicity** | | |
|  | Non-Hispanic Black | — | 1.33 (1.06, 1.66) |
|  | Hispanic | — | 2.11 (1.62, 2.75) |
|  | Other | — | 0.95 (0.42, 2.13) |
| **MCI or Dementia** | **PRIMARY EXPOSURE** | | |
|  | ***APOE* ε2 Carrier** | 0.98 (0.83, 1.15) | 0.94 (0.8, 1.11) |
|  | **DEMOGRAPHIC COVARIATES** | | |
|  | **Age** | — | 1.09 (1.09, 1.1) |
|  | **Sex - Female** | — | 0.84 (0.74, 0.95) |
|  | **Race/Ethnicity** | | |
|  | Non-Hispanic Black | — | 2.09 (1.81, 2.41) |
|  | Hispanic | — | 2.3 (1.91, 2.77) |
|  | Other | — | 1.46 (0.92, 2.3) |
| **MCI** | **PRIMARY EXPOSURE** | | |
|  | ***APOE* ε2 Carrier** | 1.04 (0.85, 1.26) | 0.99 (0.81, 1.21) |
|  | **DEMOGRAPHIC COVARIATES** | | |
|  | **Age** | — | 1.08 (1.07, 1.09) |
|  | **Sex - Female** | — | 0.78 (0.67, 0.91) |
|  | **Race/Ethnicity** | | |
|  | Non-Hispanic Black | — | 2.43 (2.05, 2.87) |
|  | Hispanic | — | 2.35 (1.87, 2.96) |
|  | Other | — | 1.57 (0.92, 2.68) |

#### eTable 16. Association Between *APOE* ε2 Carrier and Incident MCI or Dementia or Death and All-cause Mortality During the Extended Follow-up.

| **Outcome** | **Characteristic** | **Model 1**  **Unadjusted HR**  **(95% CI)** | **Model 2 Demographic-Adjusted HR**  **(95% CI)** |
| --- | --- | --- | --- |
| **MCI or Dementia or Death** | **PRIMARY EXPOSURE** | | |
|  | ***APOE* ε2 Carrier** | 0.98 (0.83, 1.15) | 0.94 (0.8, 1.11) |
|  | **DEMOGRAPHIC COVARIATES** | | |
|  | **Age** | — | 1.09 (1.09, 1.1) |
|  | **Sex - Female** | — | 0.84 (0.74, 0.95) |
|  | **Race/Ethnicity** | | |
|  | Non-Hispanic Black | — | 2.09 (1.81, 2.41) |
|  | Hispanic | — | 2.3 (1.91, 2.77) |
|  | Other | — | 1.46 (0.92, 2.3) |
| **All-cause Death** | **PRIMARY EXPOSURE** | | |
|  | ***APOE* ε2 Carrier** | 0.86 (0.76, 0.97) | 0.85 (0.75, 0.96) |
|  | **DEMOGRAPHIC COVARIATES** | | |
|  | **Age** | — | 1.08 (1.08, 1.09) |
|  | **Sex - Female** | — | 0.72 (0.65, 0.79) |
|  | **Race/Ethnicity** | | |
|  | Non-Hispanic Black | — | 1.36 (1.22, 1.51) |
|  | Hispanic | — | 0.88 (0.73, 1.05) |
|  | Other | — | 0.88 (0.6, 1.3) |

#### eTable 17. Sensitivity Analyses of Intensive vs Standard treatment effects on MCI or Dementia by *APOE* ε4 Genotype.

| **Outcome** | **Adjustment**^1^ | ***APOE* ε4** | **HR** | **UB** | **LB** | **Interaction P-value** |
| --- | --- | --- | --- | --- | --- | --- |
| Dementia | Unadjusted | Carriers | 0.73 | 0.51 | 1.04 | 0.35 |
|  |  | Non-carriers | 0.91 | 0.67 | 1.23 |  |
|  | Age | Carriers | 0.72 | 0.5 | 1.03 | 0.26 |
|  |  | Non-carriers | 0.94 | 0.7 | 1.27 |  |
|  | MoCA | Carriers | 0.77 | 0.53 | 1.1 | 0.35 |
|  |  | Non-carriers | 0.95 | 0.71 | 1.29 |  |
|  | Age & MoCA | Carriers | 0.77 | 0.54 | 1.1 | 0.21 |
|  |  | Non-carriers | 1.04 | 0.77 | 1.41 |  |
| Dementia/MCI | Unadjusted | Carriers | 0.82 | 0.65 | 1.04 | 0.60 |
|  |  | Non-carriers | 0.88 | 0.74 | 1.05 |  |
|  | Age | Carriers | 0.81 | 0.64 | 1.02 | 0.50 |
|  |  | Non-carriers | 0.89 | 0.75 | 1.06 |  |
|  | MoCA | Carriers | 0.86 | 0.68 | 1.09 | 0.81 |
|  |  | Non-carriers | 0.89 | 0.75 | 1.06 |  |
|  | Age & MoCA | Carriers | 0.87 | 0.69 | 1.1 | 0.81 |
|  |  | Non-carriers | 0.9 | 0.76 | 1.08 |  |
| MCI | Unadjusted | Carriers | 0.86 | 0.65 | 1.15 | 0.83 |
|  |  | Non-carriers | 0.83 | 0.68 | 1.01 |  |
|  | Age | Carriers | 0.84 | 0.64 | 1.12 | 0.95 |
|  |  | Non-carriers | 0.83 | 0.68 | 1.02 |  |
|  | MoCA | Carriers | 0.89 | 0.67 | 1.19 | 0.71 |
|  |  | Non-carriers | 0.84 | 0.68 | 1.02 |  |
|  | Age & MoCA | Carriers | 0.9 | 0.67 | 1.19 | 0.78 |
|  |  | Non-carriers | 0.85 | 0.7 | 1.04 |  |

^1^Adjusted models included baseline age, MoCA, and both, separately.

^2^Abbreviations: UB: upper bound; LB: lower bound. These are 95% confidence intervals.

.

#### eTable 18. Assessing Effect Heterogeneity of Intensive vs Standard SBP control by *APOE* ε4 Genotype and Race for Dementia Outcome.

| **Subgroup** | **Outcomes** | **4-year Risk** | | **RD (95% CI)** | **RR (95% CI)** | **P for interaction on RD** | **P for interaction on RR** |
| --- | --- | --- | --- | --- | --- | --- | --- |
|  |  | **Standard** | **Intensive** |  |  |  |  |
| ε4 Carriers Black | Dementia | 0.035 | 0.024 | -0.010 (-0.034, 0.013) | 0.70 (0.31, 1.59) | 0.08 | 0.14 |
|  | Dementia-free Death | 0.036 | 0.038 | 0.002 (-0.024, 0.029) | 1.06 (0.52, 2.18) |  |  |
| ε4 Carriers Non-Black | Dementia | 0.059 | 0.037 | -0.022 (-0.046, 0.001) | 0.62 (0.38, 1.03) |  |  |
|  | Dementia-free Death | 0.041 | 0.014 | -0.027 (-0.045, -0.009) | 0.35 (0.17, 0.74) |  |  |
| ε4 Non-Carriers Black | Dementia | 0.013 | 0.025 | 0.012 (-0.003, 0.027) | 1.93 (0.83, 4.48) |  |  |
|  | Dementia-free Death | 0.046 | 0.033 | -0.013 (-0.035, 0.009) | 0.72 (0.40, 1.26) |  |  |
| ε4 Non-Carriers Non-Black | Dementia | 0.022 | 0.021 | -0.001 (-0.010, 0.008) | 0.95 (0.62, 1.45) |  |  |
|  | Dementia-free Death | 0.043 | 0.029 | -0.014 (-0.026, -0.003) | 0.67 (0.48, 0.93) |  |  |

#### eTable 19. Baseline Characteristics of SPRINT and Current Study by *APOE* Genotype.

|  | **ε2 Carriers^3^** | | | **ε3 Carriers** | | | **ε4 Carriers** | | |
| --- | --- | --- | --- | --- | --- | --- | --- | --- | --- |
| **Characteristic** | **Standard**  N = 461^1^ | **Intensive**  N = 538^1^ | **SMD** | **Standard**  N = 2,250^1^ | **Intensive**  N = 2,243^1^ | **SMD** | **Standard**  N = 1,156^1^ | **Intensive**  N = 1,085^1^ | **SMD** |
| **Age** | 68.4 (9.7) | 68.4 (9.4) | 0.00 | 68.6 (9.3) | 68.5 (9.3) | 0.01 | 66.6 (9.2) | 66.8 (9.0) | -0.02 |
| **Female** | 181 (39.3%) | 208 (38.7%) | 0.60% | 778 (34.6%) | 780 (34.8%) | -0.20% | 429 (37.1%) | 403 (37.1%) | -0.03% |
| **Race** |  |  | 0.05 |  |  | 0.03 |  |  | 0.12 |
| NH White | 268 (58.1%) | 308 (57.2%) |  | 1,455 (64.7%) | 1,434 (63.9%) |  | 584 (50.5%) | 568 (52.4%) |  |
| NH Black | 157 (34.1%) | 182 (33.8%) |  | 484 (21.5%) | 481 (21.4%) |  | 452 (39.1%) | 399 (36.8%) |  |
| Hispanic | 29 (6.3%) | 40 (7.4%) |  | 271 (12.0%) | 278 (12.4%) |  | 109 (9.4%) | 93 (8.6%) |  |
| Other | 7 (1.5%) | 8 (1.5%) |  | 40 (1.8%) | 50 (2.2%) |  | 11 (1.0%) | 25 (2.3%) |  |
| **Education Level** |  |  | 0.06 |  |  | 0.04 |  |  | 0.05 |
| College graduate | 190 (41.2%) | 231 (42.9%) |  | 947 (42.1%) | 937 (41.8%) |  | 434 (37.5%) | 397 (36.6%) |  |
| High school diploma or GED | 69 (15.0%) | 85 (15.8%) |  | 340 (15.1%) | 354 (15.8%) |  | 181 (15.7%) | 189 (17.4%) |  |
| Less than high school diploma | 43 (9.3%) | 42 (7.8%) |  | 209 (9.3%) | 188 (8.4%) |  | 110 (9.5%) | 96 (8.8%) |  |
| Some college (No degree) | 159 (34.5%) | 180 (33.5%) |  | 754 (33.5%) | 764 (34.1%) |  | 431 (37.3%) | 403 (37.1%) |  |
| **Private Insurance** | 197 (42.7%) | 233 (43.3%) | -0.58% | 1,006 (44.7%) | 1,013 (45.2%) | -0.45% | 484 (41.9%) | 483 (44.5%) | -2.6% |
| **Systolic Blood Pressure** | 140.6 (15.3) | 140.1 (16.3) | 0.03 | 140.1 (15.3) | 139.6 (15.7) | 0.03 | 139.2 (15.3) | 139.7 (16.2) | -0.03 |
| **Diastolic Blood Pressure** | 78.4 (11.9) | 78.4 (12.1) | 0.00 | 77.6 (12.0) | 77.7 (11.5) | -0.01 | 78.8 (11.8) | 78.9 (12.2) | -0.01 |
| **eGFR** | 70.9 (19.7) | 72.5 (20.1) | -0.08 | 71.1 (20.1) | 71.1 (20.2) | 0.00 | 73.1 (21.1) | 72.6 (21.0) | 0.03 |
| **Glucose** | 99.1 (15.7) | 99.0 (15.0) | 0.01 | 99.3 (12.8) | 99.0 (13.4) | 0.02 | 97.5 (13.6) | 98.8 (13.6) | -0.09 |
| **Total Cholesterol** | 184.1 (38.1) | 181.9 (38.9) | 0.06 | 188.8 (40.6) | 189.3 (41.6) | -0.01 | 195.4 (41.3) | 194.8 (41.7) | 0.01 |
| **HDL Cholesterol** | 54.7 (15.6) | 54.4 (14.4) | 0.02 | 52.7 (14.2) | 52.8 (14.2) | -0.01 | 52.4 (14.7) | 51.8 (13.9) | 0.04 |
| **Statin Use** | 134 (29.1%) | 180 (33.5%) | -4.4% | 1,034 (46.0%) | 994 (44.3%) | 1.6% | 561 (48.5%) | 492 (45.3%) | 3.2% |
| **Aspirin Use** | 225 (48.8%) | 293 (54.5%) | -5.7% | 1,185 (52.7%) | 1,220 (54.4%) | -1.7% | 604 (52.2%) | 576 (53.1%) | -0.84% |
| **Smoking Status** |  |  | 0.03 |  |  | 0.01 |  |  | 0.01 |
| Current | 60 (13.0%) | 74 (13.8%) |  | 243 (10.8%) | 253 (11.3%) |  | 167 (14.5%) | 157 (14.5%) |  |
| Former | 185 (40.1%) | 210 (39.0%) |  | 997 (44.4%) | 990 (44.2%) |  | 480 (41.6%) | 446 (41.1%) |  |
| Never | 216 (46.9%) | 254 (47.2%) |  | 1,006 (44.8%) | 999 (44.6%) |  | 508 (44.0%) | 482 (44.4%) |  |
| **Body Mass Index** | 29.9 (5.7) | 30.1 (5.9) | -0.05 | 29.7 (5.6) | 29.8 (5.8) | -0.03 | 29.9 (6.0) | 29.9 (5.7) | 0.00 |
| **ACEI or ARB** | 274 (59.4%) | 316 (58.7%) | 0.70% | 1,333 (59.2%) | 1,346 (60.0%) | -0.76% | 626 (54.2%) | 633 (58.3%) | -4.2% |
| **Beta Blocker** | 149 (32.3%) | 194 (36.1%) | -3.7% | 791 (35.2%) | 851 (37.9%) | -2.8% | 399 (34.5%) | 421 (38.8%) | -4.3% |
| **Loop Diuretic** | 29 (6.3%) | 30 (5.6%) | 0.71% | 114 (5.1%) | 133 (5.9%) | -0.86% | 70 (6.1%) | 72 (6.6%) | -0.58% |
| **Calcium Channel Blocker** | 185 (40.1%) | 195 (36.2%) | 3.9% | 763 (33.9%) | 763 (34.0%) | -0.11% | 402 (34.8%) | 372 (34.3%) | 0.49% |
| **Thiazide Diuretic** | 196 (42.5%) | 206 (38.3%) | 4.2% | 926 (41.2%) | 882 (39.3%) | 1.8% | 465 (40.2%) | 440 (40.6%) | -0.33% |
| **Alpha Blocker** | 35 (7.6%) | 49 (9.1%) | -1.5% | 237 (10.5%) | 254 (11.3%) | -0.79% | 110 (9.5%) | 90 (8.3%) | 1.2% |
| **Other Antihypertensive Medication** | 15 (3.3%) | 21 (3.9%) | -0.65% | 77 (3.4%) | 88 (3.9%) | -0.50% | 51 (4.4%) | 47 (4.3%) | 0.08% |
| **Family History of Heart Disease** |  |  | 0.07 |  |  | 0.04 |  |  | 0.07 |
| No | 160 (34.7%) | 169 (31.4%) |  | 723 (32.2%) | 690 (30.8%) |  | 412 (35.6%) | 356 (32.8%) |  |
| Unknown | 25 (5.4%) | 34 (6.3%) |  | 131 (5.8%) | 119 (5.3%) |  | 61 (5.3%) | 53 (4.9%) |  |
| Yes | 276 (59.9%) | 335 (62.3%) |  | 1,392 (62.0%) | 1,434 (63.9%) |  | 683 (59.1%) | 676 (62.3%) |  |
| **Framingham Risk Score** | 20.9 (14.3, 30.9) | 20.5 (14.3, 30.1) | 0.04 | 22.5 (15.7, 32.4) | 22.5 (15.5, 32.2) | 0.01 | 21.6 (14.9, 31.3) | 21.9 (15.3, 31.5) | -0.06 |
| **MoCA Score** | 23.0 (21.0, 26.0) | 24.0 (21.0, 26.0) | -0.11 | 24.0 (21.0, 26.0) | 24.0 (21.0, 26.0) | 0.04 | 23.0 (20.0, 26.0) | 23.0 (21.0, 26.0) | -0.04 |
| **Digit Symbol Coding Test** | 50.5 (15.1) | 50.6 (14.0) | -0.01 | 51.8 (15.3) | 51.9 (15.7) | -0.01 | 51.0 (15.6) | 51.0 (14.6) | 0.00 |
| **Logical Memory Delayed Recall (LM2)** | 8.4 (3.4) | 8.3 (3.4) | 0.01 | 8.4 (3.3) | 8.4 (3.3) | 0.02 | 8.0 (3.5) | 8.3 (3.3) | -0.08 |
| ^1^Mean (SD); n (%); Median (Q1, Q3) | | | | | | | | | |
| ^2^Standardized Mean Difference; 2-sample test for equality of proportions with continuity correction  ^3^ε2 carriers = ε2/ε2 or ε3/ε2; ε3 carriers = ε3/ε3; ε4 carriers = ε4/ε2 or ε4/ε4 or ε4/ε3 | | | | | | | | | |
| Abbreviation: CI = Confidence Interval | | | | | | | | | |

#### eTable 20. Assessing Effect Heterogeneity of Intensive vs Standard SBP Control by *APOE* Genotype.

| **Outcome** | **APOE**^2^ | **Intensive Arm** | | | **Standard Arm** | | | **Intensive vs. Standard** | | | | **Interaction P-value** |
| --- | --- | --- | --- | --- | --- | --- | --- | --- | --- | --- | --- | --- |
|  |  | **N** | **Events** | **^1^Cases/**  **1,000 pys** | **N** | **Events** | **Cases/**  **1,000 pys** | **HR** | | **Lower**  **95% CL** | **Upper**  **95% CL** |  |
| Dementia | ε3/ε3 | 2243 | 76 | 6.95 | 2250 | 72 | 6.63 | 1.05 | 0.76 | | 1.45 | 0.21 |
|  | ε3/ε4 | 887 | 39 | 9.12 | 951 | 59 | 13.17 | 0.68 | 0.45 | | 1.02 |  |
|  | ε4/ε4 | 96 | 8 | 17.03 | 96 | 6 | 13.1 | 1.31 | 0.45 | | 3.76 |  |
| MCI/Dementia | ε3/ε3 | 2243 | 203 | 19.21 | 2250 | 204 | 19.53 | 0.99 | 0.82 | | 1.21 | 0.17 |
|  | ε3/ε4 | 887 | 100 | 24.38 | 951 | 134 | 31.53 | 0.78 | 0.6 | | 1.01 |  |
|  | ε4/ε4 | 96 | 17 | 38.24 | 96 | 12 | 27.46 | 1.42 | 0.68 | | 2.97 |  |
| MCI | ε3/ε3 | 2243 | 143 | 13.59 | 2250 | 158 | 15.19 | 0.90 | 0.72 | | 1.13 | 0.44 |
|  | ε3/ε4 | 887 | 72 | 17.68 | 951 | 90 | 21.4 | 0.84 | 0.62 | | 1.14 |  |
|  | ε4/ε4 | 96 | 11 | 24.88 | 96 | 9 | 20.72 | 1.22 | 0.51 | | 2.95 |  |
| MCI/Dementia/ death | ε3/ε3 | 2243 | 313 | 37.63 | 2250 | 348 | 42.01 | 0.91 | 0.78 | | 1.06 | 0.29 |
|  | ε3/ε4 | 887 | 141 | 43.26 | 951 | 183 | 53.65 | 0.75 | 0.6 | | 0.93 |  |
|  | ε4/ε4 | 96 | 19 | 53.31 | 96 | 18 | 51.75 | 1.02 | 0.54 | | 1.95 |  |
| All-cause death | ε3/ε3 | 2243 | 137 | 15.38 | 2250 | 167 | 18.83 | 0.89 | 0.71 | | 1.12 | 0.28 |
|  | ε3/ε4 | 887 | 54 | 15.21 | 951 | 62 | 16.55 | 0.77 | 0.53 | | 1.11 |  |
|  | ε4/ε4 | 96 | 3 | 7.6 | 96 | 9 | 23.27 | 0.32 | 0.09 | | 1.19 |  |
| ^1^Numbers are counts and event rates (number of cases per 1,000 person-years [pys]). The primary outcome was the first occurrence of dementia. MCI = mild cognitive impairment, CL = Confidence limit. | | | | | | | | | | | | |
| ^2^Chance imbalance was observed for several baseline covariates (age, race, education, eGFR, glucose, total cholesterol, smoking status, family history of heart disease, and logical memory delayed recall) among ε4/ε4 carriers, so the results may be subject to confounding. | | | | | | | | | | | | |

### Data Sharing Statement

**Data available**: Yes

**Data types**: Deidentified participant data, Data dictionary

**How to access data**:

- SPRINT clinical trial data: available through the NHLBI Biologic Specimen and Data Repository Information Coordinating Center (BioLINCC): <https://biolincc.nhlbi.nih.gov/studies/sprint>
- *APOE* genotype data: through the NIH National Institute on Aging Genetics of Alzheimer's Disease Data Storage Site (NIAGADS) following deposition (see "When available" for timeline)

**When available**:

- SPRINT clinical data: available beginning 07-01-2020
- *APOE* genotype data: Data will be submitted to the NIH NIAGADS within 3 months of quality control completion, per the parent NIH grant's approved data management and sharing plan. The study will also be registered in the database of Genotypes and Phenotypes (dbGaP).

**Supporting Documents / Document types**: None

**Additional Information**

**Who can access the data**: Qualified investigators with IRB/Ethics approval (or documented exemption) and a signed data use agreement, subject to the access policies of the SPRINT governing committees and, upon deposition, NIAGADS.

**Types of analyses**: General research use, biomedical use, and disease-specific use consistent with the original SPRINT informed consent, in accordance with the NIH Genomic Data Sharing Policy (NOT-OD-14-124).

**Mechanisms of data availability**: SPRINT clinical trial data through the BioLINCC data use agreement. *APOE* genotype data: interim requests may be directed to the corresponding author pending NIAGADS deposition. DNA samples remain stored at the SPRINT Central Laboratory at the University of Utah under SPRINT Steering Committee governance.
